# Causal effects on complex traits are similar across segments of different continental ancestries within admixed individuals

**DOI:** 10.1101/2022.08.16.22278868

**Authors:** Kangcheng Hou, Yi Ding, Ziqi Xu, Yue Wu, Arjun Bhattacharya, Rachel Mester, Gillian Belbin, David Conti, Burcu F. Darst, Myriam Fornage, Chris Gignoux, Xiuqing Guo, Christopher Haiman, Eimear Kenny, Michelle Kim, Charles Kooperberg, Leslie Lange, Ani Manichaikul, Kari E. North, Natalie Nudelman, Ulrike Peters, Laura J. Rasmussen-Torvik, Stephen S. Rich, Jerome I. Rotter, Heather E. Wheeler, Ying Zhou, Sriram Sankararaman, Bogdan Pasaniuc

**Affiliations:** Bioinformatics Interdepartmental Program, UCLA, Los Angeles, CA, USA; Department of Computer Science, UCLA, Los Angeles, CA, USA; Department of Pathology and Laboratory Medicine, David Geffen School of Medicine at UCLA, Los Angeles, CA, USA; Graduate Program in Biomathematics, David Geffen School of Medicine at UCLA, Los Angeles, CA, USA; Center for Genomic Health, Icahn School of Medicine at Mount Sinai, New York, NY, USA; The Charles Bronfman Institute of Personalized Medicine, Icahn School of Medicine at Mount Sinai, New York, NY, USA; Center for Genetic Epidemiology, Keck School of Medicine, University of Southern California, Los Angeles, CA, USA; Division of Public Health Science, Fred Hutchinson Cancer Center, Seattle, WA, USA; Brown Foundation Institute for Molecular Medicine, The University of Texas Health Science Center, Houston, TX, USA; Division of Biomedical Informatics and Personalized Medicine, University of Colorado, Denver, CO, USA; Institute for Translational Genomics and Population Sciences, Department of Pediatrics, Lundquist Institute at Harbor-UCLA Medical Center, Torrance, CA, USA; Department of Genetics and Genomic Sciences, Icahn School of Medicine at Mount Sinai, New York, NY, USA; Department of Medicine, Icahn School of Medicine at Mount Sinai, New York, NY, USA; Department of Medicine, University of Colorado, Aurora, CO, USA; Center for Public Health Genomics, Department of Public Health Sciences, University of Virginia, Charlottesville, VA, USA; Department of Epidemiology, University of North Carolina at Chapel Hill, Chapel Hill, NC, USA; Department of Genetics, Rutgers University, Piscataway, NJ, USA; Department of Preventive Medicine, Northwestern University Feinberg School of Medicine, Chicago, IL, USA; Department of Biology, Loyola University Chicago, Chicago, IL, USA; Program in Bioinformatics, Loyola University Chicago, Chicago, IL, USA; Department of Computational Medicine, David Geffen School of Medicine at UCLA, Los Angeles, CA, USA; Department of Human Genetics, David Geffen School of Medicine at UCLA, Los Angeles, CA, USA

**Author notes:** PAGE author names are listed in alphabetical order (indicating equal contributions).

## Abstract

Individuals of admixed ancestries (e.g., African Americans) inherit a mosaic of ancestry segments (local ancestry) originating from multiple continental ancestral populations. Their genomic diversity offers the unique opportunity of investigating genetic effects on disease across multiple ancestries within the same population. Quantifying the similarity in causal effects across local ancestries is paramount to studying genetic basis of diseases in admixed individuals. Such similarity can be defined as the genetic correlation of causal effects (*r*_admix_) across African and European local ancestry backgrounds. Existing studies investigating causal effects variability across ancestries focused on cross-continental comparisons; however, such differences could be due to heterogeneities in the definition of environment/phenotype across continental ancestries. Studying genetic effects within admixed individuals avoids these confounding factors, because the genetic effects are compared across local ancestries within the same individuals. Here, we introduce a new method that models polygenic architecture of complex traits to quantify *r*_admix_ across local ancestries. We model genome-wide causal effects that are allowed to vary by ancestry and estimate *r*_admix_ by inferring variance components of local ancestry-aware genetic relationship matrices. Our method is accurate and robust across a range of simulations. We analyze 38 complex traits in individuals of African and European admixed ancestries (*N* = 53K) from: Population Architecture using Genomics and Epidemiology (PAGE), UK Biobank (UKBB) and *All of Us* (AoU). We observe a high similarity in causal effects by ancestry in meta-analyses across traits, with estimated *r*_admix_=0.95 (95% credible interval [0.93, 0.97]), much higher than correlation in causal effects across continental ancestries. High estimated *r*_admix_ is also observed consistently for each individual trait. We replicate the high correlation in causal effects using regression-based methods from marginal GWAS summary statistics. We also report realistic scenarios where regression-based methods yield inflated estimates of heterogeneity-by-ancestry due to local ancestry-specific tagging of causal variants, and/or polygenicity. Among regression-based methods, only Deming regression is robust enough for estimation of correlation in causal effects by ancestry. In summary, causal effects on complex traits are highly similar across local ancestries and motivate genetic analyses that assume minimal heterogeneity in causal effects by ancestry.

## Introduction

Large-scale genotype-phenotype studies are increasingly analyzing diverse sets of individuals of various continental and sub-continental ancestries ^1–4^. A fundamental open question in these studies is to what extent the genetic basis of common human diseases and traits are shared/distinct across different ancestry populations ^5–9^. Understanding the role of ancestry in variability of causal effect sizes has tremendous implications for understanding the genetic basis of disease and portability of genetic risk scores in personalized and equitable genomic medicine ^1,10–13^.

The standard approach to estimating similarity in causal effects across ancestry groups has focused on cross-population analyses (typically at continental level) in which effect sizes measured by large-scale genome-wide association studies (GWAS) are compared across continental-level ancestry groups ^5–8,14,15^. Such studies have found significant differences, albeit with modest magnitude, in causal effects in cross-continental comparisons. A main drawback of such studies is the inherent differences in definition of environment/phenotype across such broad units of ancestry that can reduce the observed similarity in causal effects by ancestry; for example, the low estimated similarity in genetic causal effects for Major Depressive Disorder across Europeans and East Asians may be attributed to confounding of different diagnostic criteria in the two populations ^8,16^.

As an alternative to studying populations across different continents, causal effect similarity by ancestry can also be studied within recently admixed populations. Recently admixed individuals have the unique feature of having their genomes as mosaic of ancestry segments (*local ancestry*) originating from the ancestral populations within the past few dozen generations; for example, African American genomes are comprised of genomic segments of African and European ancestries within the past 5-15 generations ^17^. Unfortunately admixed populations are vastly under-represented in genomic studies ^18^, partly because of the lack of understanding of how the genetic causal effects vary across ancestries ^13,17,17,19–22^. For example, heterogeneity of marginal effects for a few traits and loci has been reported ^23–26^, however it remains unknown whether this reflects true difference in genetic effects or confounding due to different allele frequencies and/or linkage disequilibrium (LD) by ancestry. Recent works ^15^ have reported evidence of causal effect heterogeneity for SNPs in regions of European ancestries shared across European American and admixed African American individuals; however, they did not compare effects difference across ancestries within admixed populations. Estimating the magnitude of similarity in causal effects across ancestries is important for all genotype-phenotype studies in admixed populations from mapping to polygenic prediction, particularly within methods that allow for effects to vary across local ancestry segments ^19–22^.

In this work, we quantify the similarity in the causal effects (i.e. change in phenotype per allele substitution) across local ancestries within admixed populations; such similarity can be defined as the genetic correlation *r*_admix_ = Cor[*β* _afr_, *β* _eur_] of causal effects across African (*β* _afr_) and European (*β* _eur_) local ancestry backgrounds. We quantify *r*_admix_ using a new genetic correlation method that leverages the polygenic architecture of complex traits to include all variants (GWAS-significant and non-significant) in the model; this new approach is robust and accurate in estimating causal effects consistency across a wide range of realistic simulated genetic architectures. In addition, we also investigate regression-based approaches that use marginal effects of SNPs prioritized in GWAS risk regions. Through simulation studies, we find regression-based methods can yield deflated estimates of similarity (i.e. inflated heterogeneity) especially for highly polygenic traits and/or for studies with large differences in sample sizes across ancestries.

We analyze complex traits in African-European admixed individuals in PAGE ^1^ (24 traits, average *N* = 9K), UKBB ^2^ (26 traits, average *N* = 4K), and AoU ^3^ (10 traits, average *N* = 20K); there are 38 unique traits in total. We find causal effects are largely consistent across local ancestries within admixed individuals (through meta-analysis across 38 traits, estimated correlation of *r*_admix_ = 0.95, 95% credible interval [0.93, 0.97]). In addition, we find the heterogeneity in marginal effects exhibited at several trait-locus pairs can be explained by multiple nearby causal variants within a region, consistent with our simulation studies. Taken together, our results suggest that the causal effects are largely consistent across local ancestries within African-European admixed individuals, and this motivates future genetic analysis and method development in admixed populations that assume similar effects across ancestries for improved power.

## Results

### Overview

We start by describing the statistical model we use to relate genotype to phenotypes in two-way admixed individuals; we focus on two-way African-European admixture because their local ancestries can be accurately inferred (Methods; see Discussion for extension to other admixed populations). For a given individual, at each SNP *s*, we denote number of minor alleles from maternal and paternal haplotypes as *x*_*s*,M_, *x*_*s*,P_ ∈{0, 1} and the respective local ancestries as *γ*_*s*,M_, *γ*_*s*,P_ ∈{afr, eur}. Denoting 𝕀 (·) as the indicator function, we define the local ancestry dosage as allele counts from each of the ancestries; e.g., *𝓁*_*s*_ = 𝕀 (*γ*_*s*,M_ = afr) +𝕀 (*γ*_*s*,P_ = afr)for African (similarly for European ancestry). Conditional on genotype and local ancestry, we define ancestry-specific genotypes *g*_*s*,afr_ as the allele counts specific to the each local ancestries: *g*_*s*,afr_ := *x*_*s*,M_ 𝕀 (*γ*_*s*,M_ = afr) +*x*_*s*,P_ 𝕀 (*γ*_*s*,P_ = afr) (similarly for European ancestry, *g*_*s*,eur_). The phenotype of a given admixed individual is then modeled as function of allelic effect sizes that are allowed to vary across ancestries as:

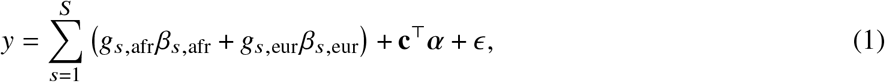

where *β*_*s*,afr_, *β*_*s*,eur_ are the causal effects at SNP *s, S* is the total number of causal SNPs in the genome, **c**, *α* denote other covariates (e.g., age, sex, genome-wide ancestries) and their corresponding effects, and *ε* is the environmental noise. *β*_*s*,afr_, *β*_*s*,eur_ are usually referred as *allelic effects*: change in phenotype with each additional allele. This is in contrast with *standardized effects* defined as change in phenotype per standard deviation increase of genotype which are usually obtained by standardizing genotype at each SNP *s* to have variance 1; i.e. 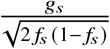 where *g* is the allele counts from all local ancestries and *f*_*s*_ is the allele frequency of SNP *s* in the population ^5,27^. We refrain from using standardized effects in this work due to the extra complexities arising from different ancestries yielding different ancestry-specific frequencies for the same SNP *s* ^5^.

Our goal is to estimate the similarity in the causal effects across local ancestries in admixed populations (Figure 1); the similarity can be evaluated across all genome-wide causal SNPs in a form of cross-ancestry genetic correlation ^5,8^: *β*_*s*,afr_, *β*_*s*,eur_ are modeled as random variable following a bi-variate Gaussian distribution parametrized by 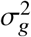, *p*_*g*_, which denote the variance and covariance of the effects, respectively:

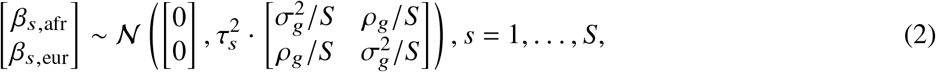

where *τ*_*s*_ are variant-specific parameters determined by the genetic architecture assumption (Methods). Under this model, the genome-wide causal effects correlation is defined as 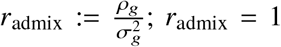 indicates same causal effects across local ancestries, while *r*_admix_ *<* 1 indicates differences across ancestries. We use a polygenic method to estimate *r*_admix_ and test the null hypothesis *H*_0_ : *r*_admix_ = 1. Specifically, given the genotype and phenotype data for a trait, we calculate the profile likelihood curve of *r*_admix_, obtained by maximizing the likelihood of model defined by Equations (1) and (2) with regard to parameters 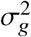 and environmental variance for each fixed *r*_admix_ ∈ [0, 1] (we assume *r*_admix_ *>* 0 a priori both because that causal effects will unlikely be negatively correlated across ancestries and to reduce *r*_admix_ search space for reducing computational cost). Then we obtain the the point estimate, credible interval and perform hypothesis testing *H*_0_ : *r*_admix_ = 1 either for each individual trait using the trait-specific profile likelihood curve, or for meta-analysis across multiple traits using the multiplication of the likelihood curves across multiple traits (analogous to inverse variance weights meta-analysis; Methods).

**Figure 1:**
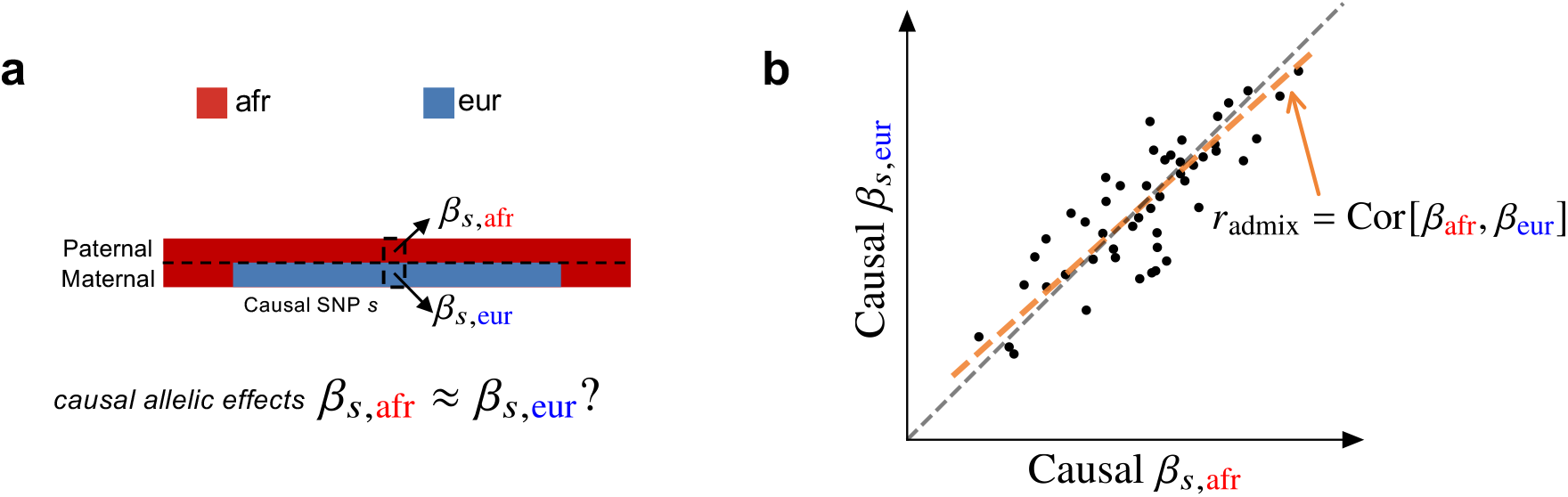
Concepts of estimating similarity in the causal effects across local ancestries. **(a)** For a given trait, with phased genotype (paternal haplotype at the top and maternal haplotype at the bottom) and inferred local ancestry (denoted by color), we investigate whether*β*_*s*,afr_ ≈ *β* _*s*,eur_ across each causal SNP *s*. **(b)** We focus on estimating the genome-wide correlation of genetic effects across ancestries *r*_admix_ = Cor[*β*_afr_, *β*_eur_], which is the regression slope (orange line) of ancestry-specific causal effects. For reference, the grey dashed line corresponds *β* _afr_ = *β* _eur_.

We organize next sections as follows. First, we show that our proposed approach provides accurate estimation of *r*_admix_ in extensive simulations. Second, we show *r*_admix_ is very close to 1 in real data of African-European admixed individuals from PAGE, UKBB and AoU. Third, we replicate our findings using methods that use GWAS summary data (i.e., marginal SNP effects at GWAS significant loci). Finally, we investigate pitfalls of methods ^4,14,15,28^ that use marginal SNP effects showing inflated heterogeneity; we find that Deming regression is the only approach robust enough to quantify *r*_admix_ from marginal GWAS effects in admixed individuals.

### Genome-wide polygenic approach to estimate genetic correlation by local ancestry is accurate in simulations

We performed simulations to evaluate our proposed polygenic method in terms of parameter estimation and hypothesis testing using real genome-wide genotypes. We simulated the phenotypes using genotypes and inferred local ancestries with *N*=17K individuals and *S*=6.9M SNPs with MAF > 0.5% in both ancestries in PAGE data set (we used these SNPs to reduce estimation variance; Methods). Phenotypes were simulated under a range of genetic architectures with a frequency dependent effects distribution for causal variants ^29,30^, varying proportion of causal variants *p*_causal_, heritability 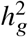, and true correlation *r*_admix_ (Methods). We used *p*_causal_ = 0.1% in our main simulation (which is close to the estimated polygenicity of a typical complex trait ^31^). When estimating *r*_admix_, we either used all SNPs in the imputed genotypes that were used to simulate phenotypes, or restricted to HapMap3 (HM3) SNPs ^32^ to simulate scenarios where causal variants are not perfectly typed in the data (Methods).

Our method produced accurate point estimates and well-calibrated credible intervals of *r*_admix_ across a wide range of realistic simulation settings (Figure 2 and tables S1 and S2). When using the imputed SNPs for estimation, results were approximately unbiased (average and maximal relative biases across simulation settings were −0.42%,-1.8% respectively). Credible intervals of *r*_admix_ meta-analyzed across simulations approximately cover true *r*_admix_: for the most biased setting 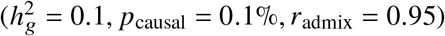, 95% credible interval = [0.915, 0.948]. When using the HM3 SNPs for estimation, there was a consistent but small downward bias (Figure 2; average and maximal relative biases were −1.0%, −2.0% respectively); correspondingly, 95% credible interval = [0.915, 0.946] for the most biased setting 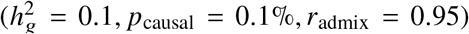. This small downward bias was due to imperfect tagging that some of the causal SNPs were not included in the HM3 SNPs. Nonetheless, the magnitude of bias from results using either imputed or HM3 SNPs was small, indicating our method was accurate and robust to imperfect tagging. We further determined our method remained accurate at other simulated *p*_causal_ (Table S2; *p*_causal_ ranging from 0.001% to 1%). Finally, in null simulations (*r*_admix_ = 1), we determined the false positive rate of our hypothesis test *H*_0_ : *r*_admix_ = 1 was properly controlled for most simulation settings, and was only slightly inflated when HM3 SNPs were used in estimation, and/or extremely low *p*_causal_ was simulated; in simulations with *r*_admix_ *<* 1, power to detect *r*_admix_ *<* 1 increased with increasing 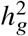 and decreasing *r*_admix_ (Tables S1 and S2). In addition, we found heritability can be accurately estimated in these simulations (Tables S3 and S4; Methods). In summary, our method can be reliably used to evaluate genome-wide genetic correlation across local ancestries (*r*_admix_).

**Figure 2:**
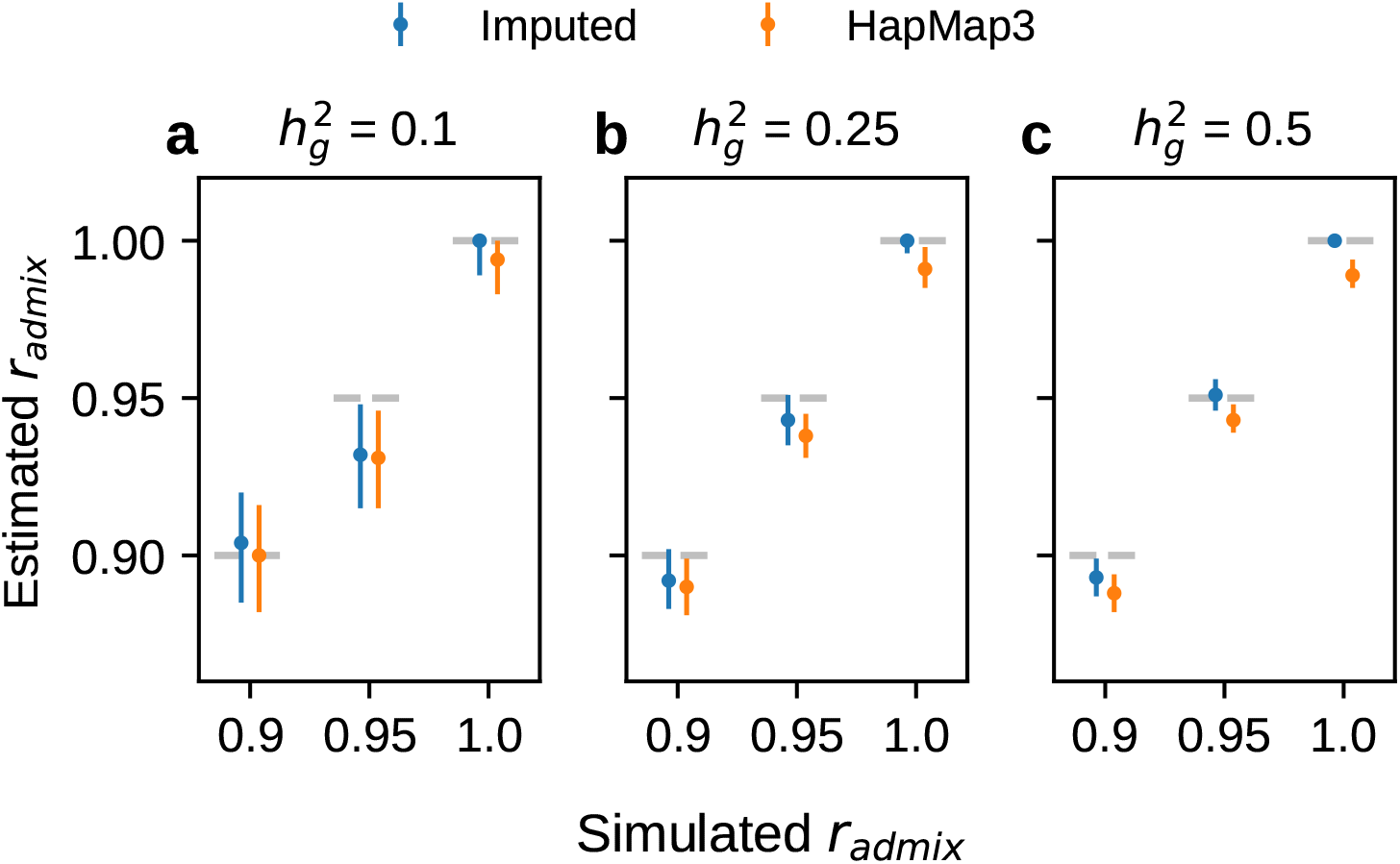
Results of genetic correlation *r*_admix_ estimation in genome-wide simulations. Simulations were based on 17K PAGE individuals and 6.9M genome-wide variants with MAF > 0.5% in both ancestries. We fixed the proportion of causal variants *p*_causal_ as 0.1%, varied genome-wide heritability 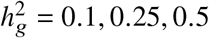, genetic correlation *r*_admix_ = 0.90, 0.95, 1.0. For each simulated genetic architecture, we plot the mode and 95% credible interval based on the meta analysis across 100 simulations (Methods). Numerical results are reported in Table S1. Numerical results for other *p*_causal_ are reported in Table S2.

### Causal effects are very similar across local ancestries in empirical data of admixed populations

We applied our polygenic method to estimate *r*_admix_ within African-European admixed individuals in PAGE ^1^ (24 traits, average *N* = 9296, average fraction of African ancestries = 78%), UKBB ^2^ (26 traits, average *N* = 3808, average fraction of African ancestries = 59%), and AoU ^3^ (10 traits, average *N* = 20496, average fraction of African ancestries = 74%) (see Methods). Meta-analyzing across 38 traits from PAGE, UKBB, AoU (60 study-trait pairs), we observed a high similarity in causal effects across ancestries (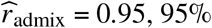 credible interval=[0.93, 0.97]). Results were highly consistent across data sets (PAGE: 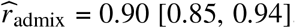, UKBB: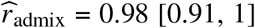, AoU: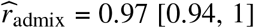) as well as traits (Figure 3a, Table 1, Table S5). Height was the only trait that had significant 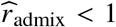 (after Bonferroni correction; nominal *p* = 4.3×10^−4^ *<* 0.05/38 meta-analyzed across three studies; Table 1) albeit with high estimated 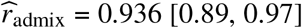. Estimates of the same traits across studies were only weakly correlated (Figure S1), suggesting similar causal effects by ancestry consistently across traits (true *r*_admix_≈1 for all traits). Our results were robust to different assumed effects distribution (Figure S2 and table S6), consistent with previous work on genetic correlation estimation ^33^. Results were also robust to the SNP set used in the estimation (Figure S2 and table S6).

**Table 1:** Genome-wide genetic correlation across 38 complex traits for African-European admixed individuals in PAGE, UKBB, AoU. For each trait, we report number of individuals, posterior mode and 95% credible interval(s) for estimated *r*_admix_, *p*-value for rejecting the null hypothesis of *H*_0_ : *r*_admix_ = 1, and estimated heritability and standard error. Meta analysis results performed across 38 traits are shown in the last row. Traits are ordered according to number of individuals. For each trait, we perform meta-analysis across studies if the trait is in multuple studies (Methods). Lymphocyte count has two credible intervals because of the non-concave profile likelihood curve, as a result of small sample size. BMD, bone mineral density. HLR, high light scattering reticulocytes. MCHC, mean corpuscular hemoglobin concentration.

| Trait | $N$ | $\hat{r}_{\text{admix}}$ mode | 95% credible interval(s) | $p$-value | $\widehat{h_g^2}$ |
| --- | --- | --- | --- | --- | --- |
| BMD | 1668 | 0.000 | [0.00, 0.78] | 0.012 | $0.34 \pm 0.16$ |
| Neuroticism | 3044 | 1.000 | [0.36, 1.00] | 1 | $0.36 \pm 0.11$ |
| Education years | 3324 | 0.000 | [0.00, 0.94] | 0.4 | $0.055 \pm 0.075$ |
| MCHC | 3650 | 0.228 | [0.00, 0.87] | 0.061 | $0.21 \pm 0.092$ |
| Type 1 diabetes | 3767 | 0.381 | [0.00, 0.95] | 0.77 | $-0.033 \pm 0.016$ |
| HLR count | 3852 | 1.000 | [0.07, 1.00] | 1 | $0.12 \pm 0.086$ |
| RBC distribution width | 3925 | 1.000 | [0.27, 1.00] | 1 | $0.28 \pm 0.087$ |
| Lymphocyte count | 3935 | 1.000 | [0.00, 0.60] [0.66, 1.00] | 1 | $0.13 \pm 0.086$ |
| Monocyte count | 3935 | 0.972 | [0.26, 1.00] | 0.82 | $0.3 \pm 0.087$ |
| MCH | 3948 | 0.829 | [0.07, 1.00] | 0.36 | $0.2 \pm 0.076$ |
| RBC count | 3948 | 1.000 | [0.37, 1.00] | 1 | $0.31 \pm 0.09$ |
| Hypothyroidism | 4063 | 1.000 | [0.05, 1.00] | 1 | $0.046 \pm 0.07$ |
| PR interval | 4071 | 0.844 | [0.08, 1.00] | 0.36 | $0.22 \pm 0.084$ |
| QRS interval | 4078 | 1.000 | [0.07, 1.00] | 1 | $0.12 \pm 0.082$ |
| Asthma | 4079 | 1.000 | [0.15, 1.00] | 1 | $0.21 \pm 0.087$ |
| Ever smoked | 4083 | 0.764 | [0.04, 0.98] | 0.31 | $0.17 \pm 0.082$ |
| QT interval | 4089 | 0.920 | [0.07, 1.00] | 0.69 | $0.16 \pm 0.083$ |
| HbA1c | 5353 | 0.954 | [0.08, 1.00] | 0.77 | $0.19 \pm 0.078$ |
| Cigarettes per day | 6995 | 0.999 | [0.08, 1.00] | 1 | $0.097 \pm 0.047$ |
| Fasting insulin | 7753 | 1.000 | [0.21, 1.00] | 1 | $0.13 \pm 0.044$ |
| eGFR | 7978 | 0.805 | [0.16, 1.00] | 0.09 | $0.19 \pm 0.046$ |
| C-reactive protein | 8321 | 0.995 | [0.82, 1.00] | 0.94 | $0.28 \pm 0.046$ |
| Fasting glucose | 9646 | 0.695 | [0.00, 0.93] | 0.27 | $0.064 \pm 0.035$ |
| Coffee consumption | 11587 | 0.982 | [0.10, 1.00] | 0.9 | $0.074 \pm 0.03$ |
| Platelet count | 12545 | 0.783 | [0.20, 0.98] | 0.025 | $0.19 \pm 0.038$ |
| White blood cell count | 12755 | 0.931 | [0.70, 1.00] | 0.26 | $0.23 \pm 0.036$ |
| Type 2 diabetes | 18630 | 0.897 | [0.49, 1.00] | 0.23 | $0.12 \pm 0.024$ |
| Hypertension | 20744 | 0.929 | [0.30, 1.00] | 0.45 | $0.08 \pm 0.027$ |
| LDL | 21979 | 0.958 | [0.70, 1.00] | 0.55 | $0.14 \pm 0.046$ |
| HDL | 22039 | 0.961 | [0.82, 1.00] | 0.46 | $0.22 \pm 0.057$ |
| Triglycerides | 22494 | 0.843 | [0.54, 0.98] | 0.012 | $0.18 \pm 0.027$ |
| Total cholesterol | 22555 | 0.818 | [0.50, 0.97] | 0.007 | $0.18 \pm 0.039$ |
| Heart rate | 28764 | 0.980 | [0.82, 1.00] | 0.74 | $0.099 \pm 0.015$ |
| WHR | 36756 | 0.973 | [0.86, 1.00] | 0.55 | $0.12 \pm 0.015$ |
| Diastolic blood pressure | 43787 | 1.000 | [0.90, 1.00] | 1 | $0.077 \pm 0.024$ |
| Systolic blood pressure | 43788 | 1.000 | [0.88, 1.00] | 1 | $0.071 \pm 0.013$ |
| BMI | 49521 | 0.974 | [0.92, 1.00] | 0.33 | $0.22 \pm 0.02$ |
| Height | 49605 | 0.936 | [0.89, 0.97] | 0.00043 | $0.4 \pm 0.014$ |
| Meta analysis | | 0.947 | [0.93, 0.97] | $8.7 \times 10^{-7}$ | |

**Figure 3:**
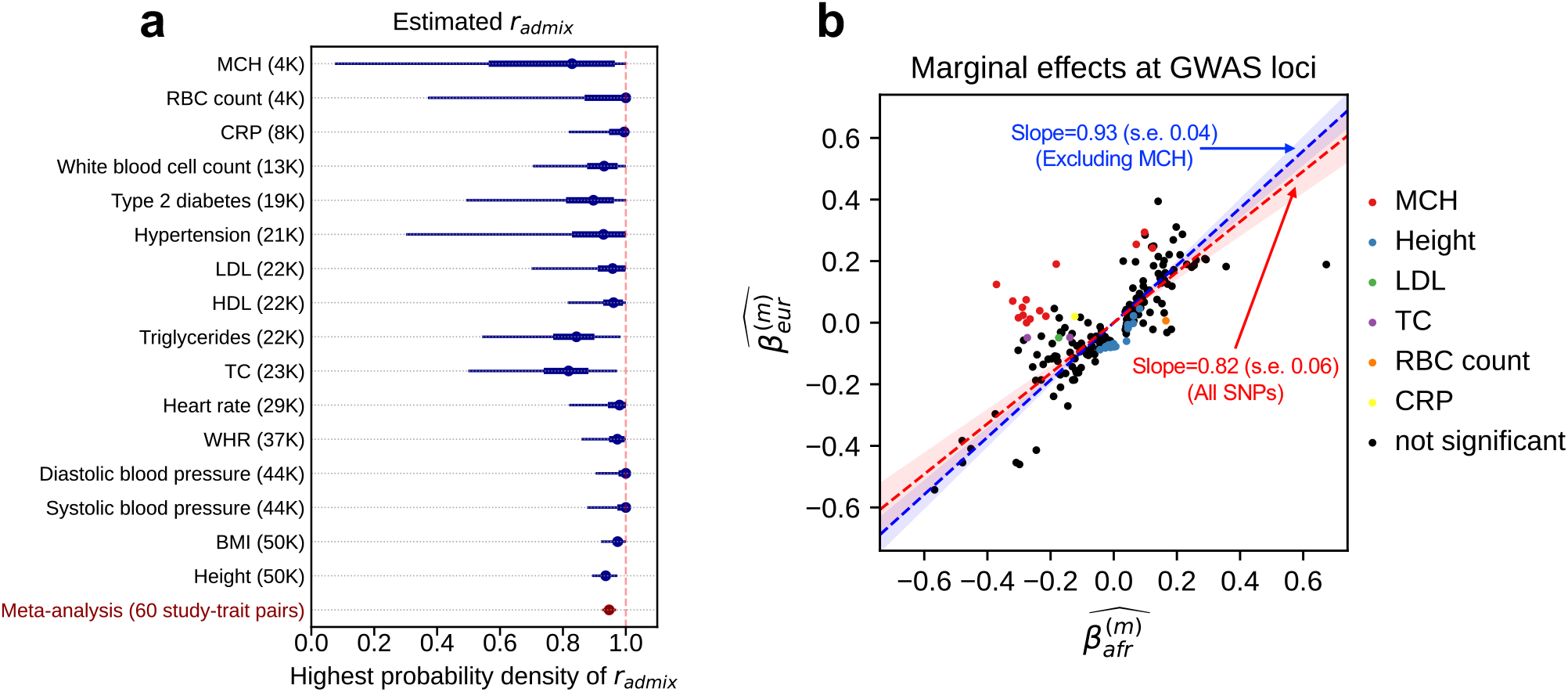
Similarity of causal effects and marginal effects across local ancestries meta-analyzed across PAGE, UKBB, AoU. **(a)** We plot the trait-specific estimated *r*_admix_ for 16 traits. For each trait, dots denote the estimation modes; bold lines and thin lines denote 50% / 95% highest density credible intervals, respectively. Traits are ordered according to total number of individuals included in the estimation (shown in parentheses). These traits are selected to be displayed either because they have the largest total sample sizes, or because the associated SNPs of these traits exhibit heterogeneity in marginal effects (see the panel on the right). We also display the meta-analysis results across all 38 traits (60 study-trait pairs). **(b)** We plot the ancestry-specific marginal effects for 217 GWAS significant clumped trait-SNP pairs across 60 study-trait pairs. Trait-SNP pairs with significant heterogeneity in marginal effects by ancestry (*p*_HET_ *<* 0.05/217 via HET test) are denoted in color (non-significant trait-SNP pairs denoted as black dots). Numerical results are reported in Table S7. Deming regression slopes of 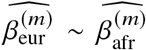 are provided either for all 217 SNPs, or for 193 SNPs after excluding 24 MCH-associated SNPs. MCH, mean corpuscular hemoglobin. RBC, red blood cell. CRP, C-reactive protein. LDL, low density lipoprotein cholesterol. HDL, high density lipoprotein cholesterol. TC, total cholesterol. BMI, body mass index. WHR, waist to hip ratio. Numerical results are provided in Tables 1 and S7.

Next, we contrasted *r*_admix_ to trans-continental genetic correlations (between Europeans and East Asians) ^8^. We found a larger similarity across local ancestries within admixed populations as opposed to trans-continental correlations: 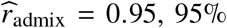 credible interval [0.93, 0.97] vs. 0.85, 95% confidence interval [0.83, 0.87] ^8^. Although the traits considered in these studies only partially overlap, our results are consistent with differences in phenotyping/environment across continents reducing the observed genetic correlations in trans-continental studies.

We sought to replicate high *r*_admix_ using regression-based methods that leverage estimated ancestry-specific marginal effects at GWAS loci (Methods). Specifically, we use the following marginal regression equation (restricting Equation (1) to each GWAS-clumped SNP *s*): 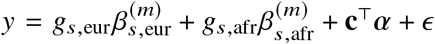 (we distinguish marginal effects *β*^(*m*)^ from causal effects *β*; Methods). Across 60 study-trait pairs, we detected a total of 217 GWAS significant clumped trait-SNP pairs and we estimated the ancestry-specific marginal effects for each of these SNPs (Figure 3b, Table S7). We determined the estimated marginal effects are largely consistent by local ancestry at these GWAS clumped SNPs via Deming regression slope ^34^ of 0.82 (SE 0.06) (applied to 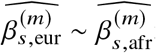 Methods). Mean corpuscular hemoglobin (MCH)-associated SNPs at 16p13.3 drove the most of the differences by ancestry: Deming regression slope was 0.93 (SE 0.04) on the rest of 193 SNPs after excluding 24 MCH-associated SNPs; MCH-associated SNPs also have the strongest heterogeneity in marginal effects by ancestry (using HET test, an 1-degree of freedom test for allelic effects heterogeneity at each SNP ^35^; Table S7; Methods). We found there are multiple conditionally independent association signals at MCH-associated loci (Figure S3) and other loci (Figures S4 and S5) that had heterogeneity by ancestry by performing statistical fine-mapping analysis (Methods; Supplementary Notes). In fact, the MCH-associated loci locate at a region harboring alpha-globin gene cluster (*HBZ-HBM-HBA2-HBA1-HBQ1*) known to harbor multiple causal variants ^36^. These results suggest that, similar to causal effects, marginal effects at GWAS loci are also largely consistent by local ancestry, except that loci with multiple causal variants can drive some extent of heterogeneity by ancestry in marginal effects.

### Pitfalls of using marginal effects at GWAS significant variants to estimate heterogeneity in causal effects

Next, we focused on thoroughly evaluating methods that use marginal effects at GWAS significant variants to estimate genetic correlation. Marginal effects are frequently used to compare effect sizes across populations or across studies ^4,14,15,28^ and enjoy great popularity for their simplicity and requirement of only GWAS summary statistics (estimated effect sizes and standard errors).

We first note that the heterogeneities in marginal effects can be induced due to different LD patterns across ancestries even when the underlying causal effects are identical, especially when multiple causal variants are nearby in the same LD block (Figure 4). We investigate the extent of heterogeneity by ancestry that can be induced in simulations with identical causal effects across ancestries, due to (1) local ancestry adjustment; (2) unknown causal variants coupled with ancestry specific LD patterns; (3) highly polygenic trait architectures with multiple causal SNPs within the same LD block; (4) differential GWAS sample sizes across ancestries. Our following simulations were based on real imputed genotypes from African-European individuals in PAGE data (17K individuals, average fraction of African ancestries = 78%).

**Figure 4:**
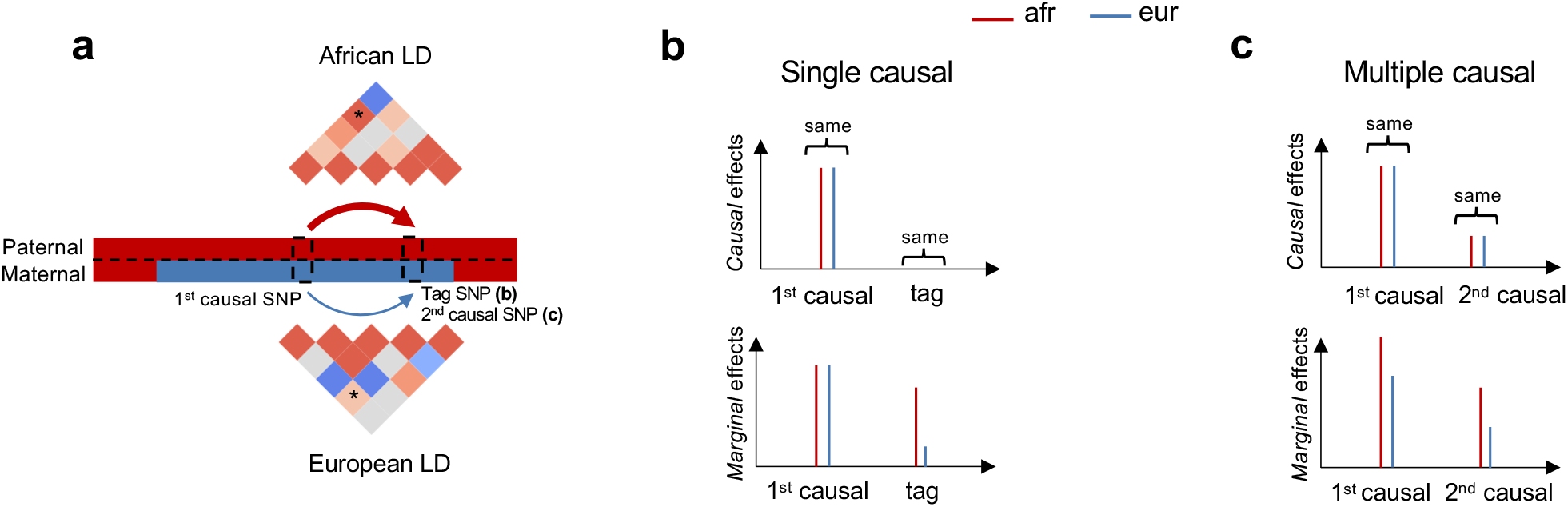
Induced heterogeneities in marginal effects across local ancestries. **(a)** Illustrations that different LD patterns across local ancestries can induce differential tagging between a causal SNP and a tag SNP in **(b)** or another causal SNP in **(c)**. LD strengths between the two SNPs are indicated both in the thickness of arrows and in the color shades of ‘*’ elements in LD matrices. **(b)** Example of single causal SNP with no heterogeneity. Causal effects are the same across local ancestries, and the estimated marginal effects at causal SNP will be also very similar with sufficient sample size. However, because of differential tagging across local ancestries, the estimated marginal effects evaluated at the tag SNP are different. **(c)** Example of multiple causal SNPs with no heterogeneity. Causal effects for both SNPs are the same across local ancestries. In this example, the correlation between the 2 causal variants is higher for genotypes in African local ancestries than those in European local ancestries. Therefore, African ancestry-specific genotypes tag more effects, creating different marginal ancestry-specific effects at each causal SNP.

### Adjusting for local ancestry can deflate the observed similarity in causal effects across ancestries

We first discuss the use of local ancestry in the heterogeneity estimation, which is a unique and important component to consider when studying admixed populations. We used simulations to investigate the role of local ancestry adjustment in heterogeneity estimation using three main approaches: (1) ignoring local ancestry altogether (“w/o”); (2) including local ancestry as covariate in the model (“lanc-included”); (3) regressing out the local ancestry from phenotype followed by heterogeneity estimation on residuals (“lanc-regressed”) (Methods). First, in null simulations where causal effects are similar across ancestries (i.e. ratio of *β*_eur_ to *β*_afr_ = 1), we observed that strategies of ignoring local ancestry altogether or including a covariate for local ancestry in the model yielded well-calibrated HET tests; in contrast, the approach of regressing out the local ancestry effect prior to assessing heterogeneity induced inflated HET test statistics (Figure 5 and table S8). Next, we used power simulations where we varied the amount of heterogeneity (defined as ratio of *β*_eur_ : *β*_afr_): as expected, including local ancestry in the covariate significantly reduced the power of HET test of up to 50% at high magnitude of heterogeneity (Figure 5 and table S8) (detailed explanation of these observations can be found in Supplementary Notes). Thus, with respect to local ancestry, we recommend either not using it or including it as a covariate in the model and not regressing out its effect prior to heterogeneity estimation as that will bias heterogeneity estimation.

**Figure 5:**
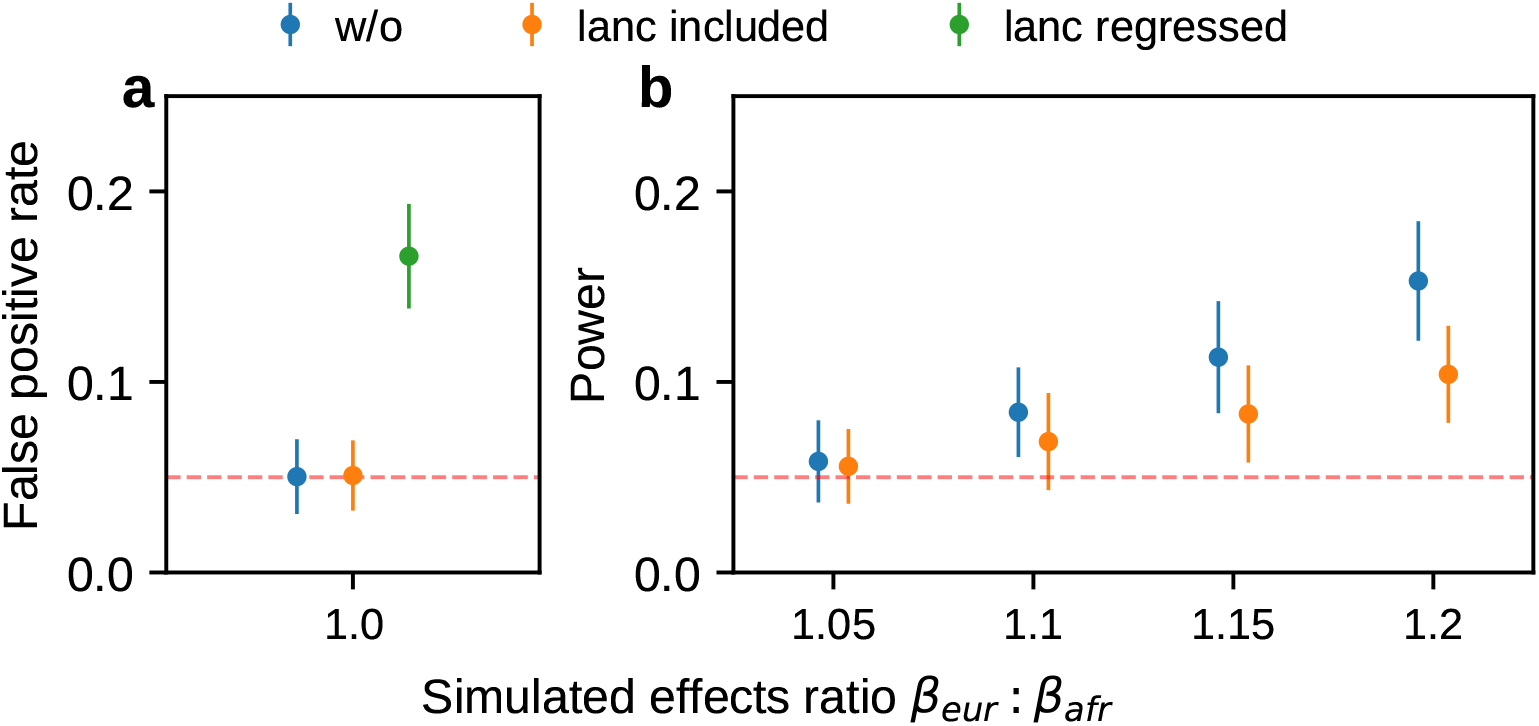
Pitfalls of including local ancestry in estimating heterogeneity. In each simulation, we selected a single causal variant and simulated quantitative phenotypes where these causal variants explain heritability 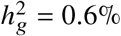; we also varied ratios of effects across ancestries *β*_eur_ : *β*_afr_. **(a)** False positive rate in null simulation *β*_eur_ : *β*_afr_ = 1.0. (b) Power to detect *β*_eur_ ≠ *β*_afr_ in power simulations with *β*_eur_ : *β*_afr_ *>* 1. 95% confidence intervals are calculated based on 100 random sub-samplings with each sample consisting of 500 SNPs (Methods). Numerical results are reported in Table S8.

Having investigated the role of local ancestry adjustment, we next turn to heterogeneity estimation for GWAS clumped SNPs. We investigated properties of HET test and Deming regression in simulations with *r*_admix_ = 1. Since the true causal variants are unknown and need to be inferred, we investigated each method either at the true simulated causal variants or at the clumped variants from LD clumping (Methods).

### Unknown causal variants can deflate the observed similarity in effects by ancestry

We first performed simulations with single causal variant: in each simulation, we randomly selected 1 SNP as causal. Evaluated at the simulated causal SNPs (Methods), we found that HET test and Deming slope were well-calibrated (Figure 6 and table S9). However, evaluated at the clumped variants, as a more realistic setting (because causal variants need to be inferred), we found HET test became increasingly mis-calibrated with increased 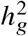, while Deming slope remained relatively robust (with an upward trend although not statistically significant with increasing 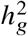) (Figure 6ab).

**Figure 6:**
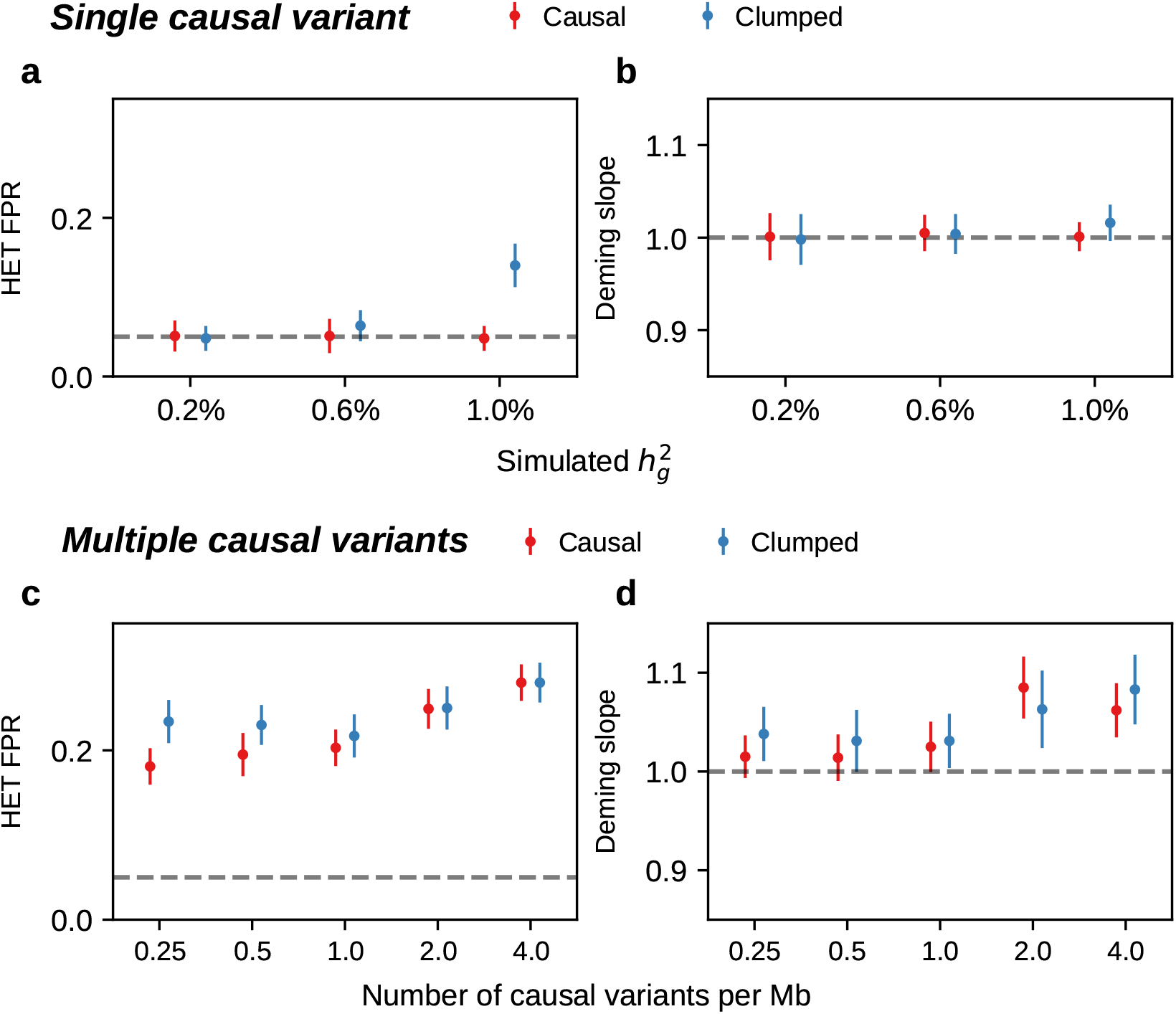
aMis-calibration of HET test and Deming regression slope in simulations with *r*_admix_ = 1. **(a-b)** Simulations with single causal variant. Each causal variant had the same causal effects across local ancestries and each causal variant explained a fixed amount of heritability (0.2%, 0.6%, 1.0%). **(a)** False positive rate (FPR) of HET test. **(b)** Deming regression slope of 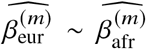. Numerical results are reported in Table S9. **(c-d)** Simulation with multiple causal variants. We simulated different level of polygenicity, such that on average there were approximately 0.25, 0.5, 1.0, 2.0, 4.0 causal variants per Mb. Causal variants had same causal effects across local ancestries. The heritability explained by all causal variants was fixed at 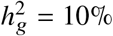. **(c)** FPR of HET test. **(d)** Deming regression slope of 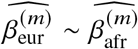. 95% confidence intervals were based on 100 random sub-samplings with each sample consists of 1,000 SNPs (Methods). Results for other number of SNPs used for sub-sampling are shown in Figure S7. Numerical results are reported in Table S10.

### High polygenicity can deflate the observed similarity in effects by ancestry

Next, we turn to simulations where multiple causal variants are likely to occur within the same LD block (typical for complex polygenic traits ^37,38^; Methods). In this scenario, marginal GWAS effects could tag multiple causal effects thus potentially deflating the observed heterogeneity (Figure 4c). In simulations, we varied the number of causal SNPs from 0.25 to 4.0 per Mb to span most polygenic architectures. In contrast to the scenario of a single causal variant, both HET test and Deming slope were biased in the presence of multiple causals within the same LD region; the mis-calibration/bias increased with number of causals per region (Figure 6cd). LD clumping did not alleviate the mis-calibration/bias (Figure 6cd). Such mis-calibrations occurred irrespective of sample size (Figure S7), or simulated heritability 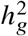 (Table S10).

In summary, we find that methods for heterogeneity-by-ancestry estimation based on marginal GWAS SNP effects are susceptible to inflated estimates of heterogeneity. HET test is susceptible to false positives when causal variants are unknown. Deming regression was robust in scenarios with low polygenicity, however, was susceptible to inflated estimates of heterogeneity for simulated highly polygenic traits; the inflated estimates can be explained by differential tagging by ancestry of causal effects across ancestries among causal SNPs. We also investigated other regression-based methods, including ordinary least squares (OLS) slope and Pearson’s correlation. Compared to Deming regression slope, we determined they are even more biased due to their ignorance of the errors in the estimated effects, especially in the presence of differential GWAS sample sizes across local ancestries. We provide detailed discussions in Methods and Supplementary Notes.

## Discussion

In this work, we developed a new polygenic method that model genome-wide causal effects to complex traits of admixed individuals and applied our method to 53K African-European admixed individuals across 38 complex traits in PAGE, UKBB, AoU. We determined causal effects are largely consistent across local ancestries. In addition to causal effects, we also replicated such consistency-by-ancestry for marginal effects at GWAS loci. We highlighted realistic simulation scenarios where regression-based methods using marginal effects can report false heterogeneity when causal effects are identical across ancestries.

Our study has several implications for future genetic study of admixed populations. First, reduced accuracy of polygenic score has been observed in African-European admixed populations with increasing proportion of non-European ancestries, and it was previously unknown on the relative contribution of causal effects difference to this reduced accuracy, among other factors such as MAF and LD difference across ancestries ^21^. Our results suggest the causal effects difference has limited contribution to this reduced accuracy. Second, there have been recent work on incorporating local ancestry in statistical modeling of admixed populations, e.g. in association testing ^19^, polygenic score ^21,22^, based on the hypothesis that effects may differ across ancestries. Our results indicate the largely consistent causal effects across local ancestries (and also marginal effects at most GWAS loci). The robustness of our results to imperfect tagging (in simulation and real data analyses using HM3 SNPs) also suggests that imperfect tagging induce limited effects heterogeneity across local ancestries, once SNPs are properly modeled in a polygenic model. Therefore, our results suggest future genetic analysis within admixed individuals should prioritize statistical models assuming same causal effects across local ancestries for improved statistical power.

Our results add to the existing literature to further delineate sources of causal effects differences. Previous works demonstrated moderate causal effects differences across trans-ethnic populations ^5,6,8^. Similarly, a recent work ^15^ concluded differences between causal effects in European local ancestries within African American admixed individuals and that in European American individuals. Our results indicate more similar causal effects across local ancestries than across trans-continental populations, which is likely explained by the absence of gene-environment interaction differences across local ancestries. On the other hand, the small differences across local ancestries, if exist, may be attributed to the local genetic interaction.

We note several limitations and future directions of our work. First, we have analyzed SNPs with MAF > 0.5% in both ancestries. We excluded population-specific SNPs (with MAF≤0.5% in one of the ancestries) because they provide little information for estimating the consistency of causal effects, since the effects are estimated with large noises for these SNPs. We hypothesize that, if any, the exclusion of these population-specific SNPs can produce a downward bias of estimated genetic correlation, similar to the bias in our simulation with HapMap3 SNPs. Therefore, the estimated genetic correlation here are likely a lower bound of the true genetic correlation. Second, we have considered two-way African-European admixed individuals. Several practical considerations remain before applying this method to other admixed populations such as three-way admixture of Latino American populations: local ancestries are typically inferred with larger errors ^39^ and should be accounted for in statistical modeling, and additional parameters need to be estimated (e.g., three pairwise correlation parameters across ancestries for three-way admixture populations). In addition, for Latino American populations, given the large noises in estimated African local ancestries because of their small proportion ^40^, it may be desired to alternatively estimate genetic correlation of Native American ancestries vs. other ancestries (including both European and African ancestries). In this case, the estimated genetic correlation can be interpreted as differences of causal effects in Native American local ancestries versus the average causal effects of European and African local ancestries. We leave extension to other admixed populations for future work. Third, we have focused on estimating a single global parameter *r*_admix_ which summarizes the overall genome-wide genetic correlation. Our modeling framework can be extended to stratified analyses of SNPs in different annotation categories (e.g., MAF bins or functional annotations ^41^) to estimate the genetic correlation within each category. To obtain estimates with sufficient precision for each SNP category, such stratified analyses would require larger sample sizes compared to the overall analyses we performed here. We leave such stratified analyses for future work with access to larger sample sizes of admixed individuals. Fourth, our polygenic method requires individual-level genotype and phenotype; if not available, we found Deming regression may be applied to evaluate heterogeneity with caution: in our simulation, Deming regression was the only method robust to most scenarios except for high polygenicity. Fifth, although we have meta-analyzed three publicly available studies with large cohort of African-European admixed individuals, our estimate for each individual trait was still associated with large standard errors and can be further improved by analyzing more individuals. Sixth, methods described here can be readily applied to gene expression data of admixed individuals to investigate heterogeneity for gene expression; we leave this to future work because such data with large sample size is currently unavailable to us.

Despite these limitations, our study has shown that causal effects to complex traits are highly similar across local ancestries within European-African admixed populations and this knowledge can be used to guide future genetic studies of admixed populations.

## Methods

### Statistical model of phenotype for admixed individuals

For each individual *i* = 1, …, *N* and SNP *s* = 1, …, *S*, we denote *x*_*i,s*,M_, *x*_*i,s*,P_ as number of minor alleles at maternal haplotype and paternal haplotype, respectively. We assume the corresponding local ancestries *γ*_*i,s*,M_, *γ*_*i,s*,P_ ∈{1, 2} are inferred accurately (we focus on 2-way admixed populations in this work). For example, for African-European admixed individuals, ‘1’ corresponds to African ancestries, ‘2’ corresponds to European ancestries. For each individual *i* and SNP *s*, we define ancestry-specific genotypes, *g*_*i,s*,1_ for ancestry 1 and *g*_*i,s*,2_ for ancestry 2, as the allele counts that are specific to each local ancestry:

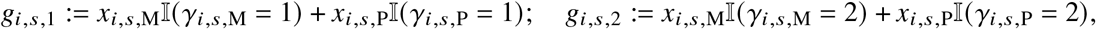

where 𝕀(·) denotes the indicator function. Denoting the causal allelic effects as ***β***_1_, ***β***_2_ ∈ ℝ^*S*^ for the two ancestries, we model the phenotype of each individual *y*_*i*_ as

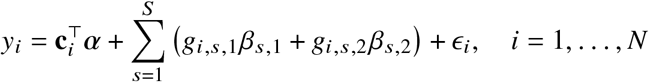

where **c**_*i*_ ∈ ℝ^*C*^, ***α*** ∈ ℝ^*C*^ are the *C* individual-specific covariates (including the all ‘1’ intercepts) and the corresponding effects. *ϵ*_*i*_ is the i.i.d. environmental factors for each individual *i*. If we further denote the ancestry-specific genotype matrix as **G**_1_∈{0, 1, 2} ^*N*×*S*^ and **G**_2_∈{0, 1, 2} ^*N*×*S*^for ancestry 1 and 2, and **C** ∈ ℝ^*N*×*S*^ as the covariates matrix, we can write Equation (1) as

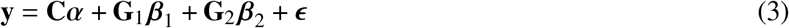

We pose the following distribution assumptions on the effect sizes and environmental noises

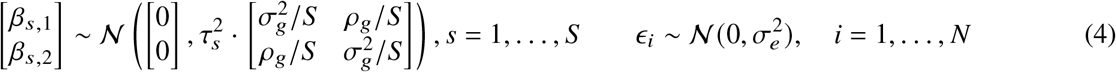

where 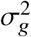 is the variance of effect sizes for the two populations, *ρ*_*g*_ is the covariance parameter measuring the similarity of effect sizes from the two populations, and 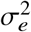 is the variance parameter for the environmental factors. *τ*_*s*_ are SNP-specific parameters (estimated and fixed a priori) for specifying the effect sizes distribution (see “Specifying *τ*_*s*_ under different heritability models” below). We define the genome-wide genetic correlation as 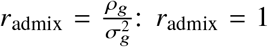 indicates *β*_*s*,1_ =*β*_*s*,2_ for all variants *s* = 1, …, *S*, i.e., causal allelic effects sizes are the same across ancestries; *r*_admix_ *<* 1 indicates some level of differences in causal allelic effect sizes across ancestries.

### Calculating and filtering by local ancestry-specific allele frequencies

For each SNP *s*, we calculated the minor allele frequency with 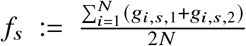. We also calculated the ancestry-specific allele frequency as 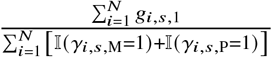 for ancestry 1, and similarly 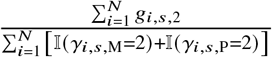 for ancestry 2. For a SNP *s* with zero frequency (or very low frequency) for either of the ancestry, the corresponding effect size *β*_*s*_ will be unidentifiable (or estimated with very large noise). Therefore, we filtered for SNPs with MAF > 0.5% for both ancestries throughout in analyses.

### Defining heritability

Under the above assumptions, the heritability 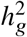 can be derived as a function of 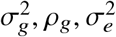 as general form of 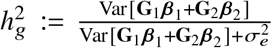. Suppose *r*_admix_ = 1, or equivalently 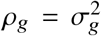, since *β*_1_ and *β*_2_ is now perfectly correlated, we denote *β* = *β*_1_ = *β*_2_ as the per-SNP effect sizes. Defining 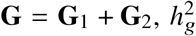 can be written as 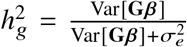. Assuming genotype matrix **G** is centered for each SNP, 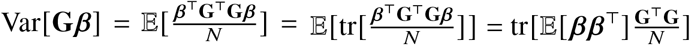. Noting that E[*β β* ^T^] is a diagonal matrix with *s*-th element being 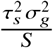 and *s*-th element of 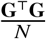 diagonal being SNP variance 2 *f*_*s*_ (1 − *f*_*s*_), Var[**G***β*] can be calculated with

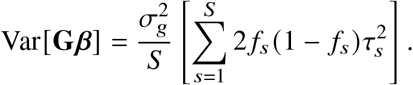

We have made the assumption of *r*_admix_ = 1 when deriving the heritability formula, and we have shown through simulations that this formula leads to accurate estimates of heritability in simulation even when this assumption is violated (with *r*_admix_ = 0.9, 0.95, 1.0; Tables S3 and S4). Given the simplicity of this formula and that our real data analysis results indicate *r*_admix_ *>* 0.9 (Table 1), we used this formula throughout in this work. In our real data analysis, each trait can have heritability estimates from multiple studies. To obtain a single estimate of heritability for each trait, we performed random-effects meta-analysis using point estimates and standard error of the heritability obtained in each study (i.e. heritability estimates in Table 1 are per-trait meta-analysis results from heritability estimates in Table S5).

### Specifying *τ*_*s*_ under different heritability models

*τ*_*s*_ can be used to specify the coupling of SNP effects variance with MAF, local LD or other functional annotations. Commonly used heritability models include

- GCTA model ^42^: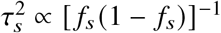, where *f*_*s*_ is the MAF of the SNP *s*.
- Frequency-dependent model ^29,30^: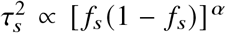, where *α* specifies the coupling between per-allele effect sizes and MAF of SNP *s*. We note that when *α* = −1, this becomes the GCTA model.
- LDAK ^33^: 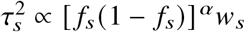, where *w*_*s*_ are the SNP-specific LDAK weights, as a function of the inverse of the local LD of SNP *s*. The parameter *α* controls the coupling between SNP effects and SNP frequency.
- S-LDSC ^43^: 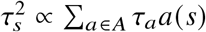, where each *a∈A* corresponds to a set of binary or continuous annotations representing MAF, local LD or other functional annotations. *a(s)* is the value of the annotation *α* for SNP *s*, and *τ*_*a*_ corresponds to the expected increase of 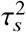 with each additional unit of annotation *α*.

Choice of heritability model has shown to be important to study heritability and functional enrichment of heritability ^33,44,45^. However, genetic correlation estimation, the main focus of this study, has shown to be robust to different heritability model ^33^. In this work, we mainly used the frequency-dependent model with *α* = −0.38 (from a previous meta-analysis across 25 UK Biobank complex traits ^30^) for both simulation and estimation. For real data analysis, we additionally used GCTA model for estimation and found results are robust to heritability models (Figure S2), consistent with previous study ^33^.

### Evaluation of genome-wide genetic effects consistency

We discuss parameter estimation and hypothesis testing in Equations (3) and (4). First, we note that, marginalizing over random effects *β* _1_ and *β* _2_ in Equation (3), the distribution of **y** is

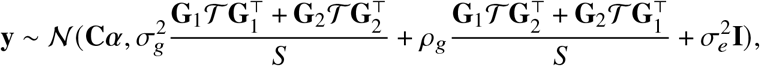

where *𝒯* is a diagonal matrix with 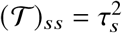. We note that 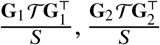 can be thought as local ancestry-specific genetic relationship matrices. Denoting that 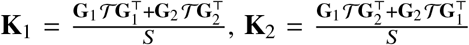, and 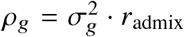, the distribution of **y** can be simplified as

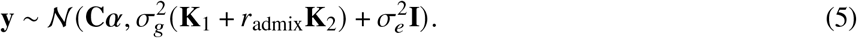

The maximum likelihood estimate of 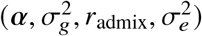 can be found by directly maximizing the corresponding likelihood function 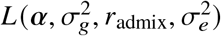. Ho wever, by definition of correlation, *r*_admix_ should satisfy the constraint of *r*_admix_ ≤ 1. And it is not straightforward to put this constraint into optimization. Therefore, we use the profile likelihood 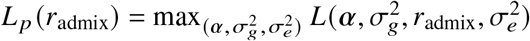 and perform grid search of *r*_admix_ to maximize this profile likelihood (similar to ref.): for each candidate *r*_admix_, we compute **K**_1_ + *r*_admix_**K**_2_, and solve 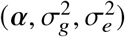 for the single variance component model as defined in Equation (5) using AI-REML implemented in GCTA software ^27^. In practice, we calculate profile likelihood *L*_*p*_(*r*_admix_) for a predefined set of *r*_admix_ = 0.00, 0.05, …, 1.00 (we note that *r*_admix_ ∈[0, 1] is a reasonable prior assumption in our work; this predefined set can be extended to other range), and use natural cubic spline to interpolate pairs of (*r*_admix_, *L*_*p*_(*r* _admix_)) to get a likelihood curve of *r* _admix_. Then we obtain the estimated 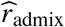 using the value that maximize the likelihood curve, and credible interval by combining the likelihood curve with a uniform prior of *r*_admix_∼Uniform [0, 1]and calculating the highest posterior density interval. To perform the meta-analysis across independent estimates, we first obtain the joint likelihood by calculating the product of likelihood curves across estimates (or equivalently, the sum of log-likelihood curves), and then calculate the estimate and credible interval same as described above.

### Evaluation of genetic effects consistency at individual variant with marginal effects

#### Parameter estimation and hypothesis testing

We construct a model between individual variant *s* and phenotype by restricting Equation (1) to the specific SNP of interest *s*, as

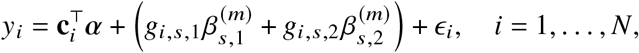

or in vector form,

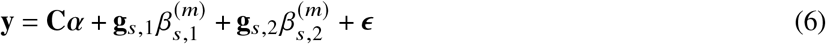

where **C, g**_*s*,1_, **g**_*s*,2_, ***ϵ*** contain **c**_*i*_, *g*_*i,s*,1_, *g*_*i,s*,2_, ***ϵ***_*i*_ for all individuals *i* = 1, …, *N*, respectively. We distinguish the marginal effects 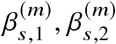 in Equation (6) from causal effects *β*_*s*,1_, *β*_*s*,2_ in Equation (1): marginal effects at the GWAS-clumped SNP tag effects from nearby causal SNPs with taggability as a function of ancestry-specific correlation between the GWAS-clumped SNP and nearby causal SNPs, and therefore, heterogeneity in marginal effects by local ancestry can be induced even if causal effects are the same (see extensive simulation studies above).

We estimate effect sizes for two ancestries 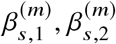 using least squares (jointly for 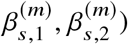) and perform hypothesis testing of 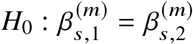 with a likelihood ratio test by comparing Equation (6) to a restricted model where the allelic effects are the same 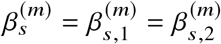:

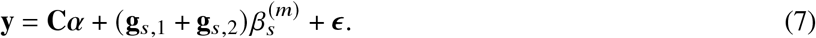

Twice the difference of log-likelihood follows a chi-square distribution with degree of freedom 1. We note that this procedure is similar to ref. ^19^ but we do not include local ancestry in the model.

#### Induced marginal effects heterogeneity due to tagging

We describe the induced heterogeneity of estimated marginal effects at tagging variants, even when causal variant effects are the same across ancestries. We consider two variants *s, t*, with variant *s* as the causal variant and variant *t* as the tagging variant. For simplicity, we ignore the covariates and assume **y, g**_*s*,1_, **g**_*s*,2_ have been centered (equivalent to including the all ‘1’ covariate in the model); similar results can be derived for scenarios with covariates, by projecting **y, g**_*s*,1_, **g**_*s*,2_ out of the covariate space.

We first assume *s* as the only causal variant, therefore, phenotype can be modeled as **y** = **g**_*s*,1_ *β*_*s*,1_ +**g**_*s*,2_*β*_*s*,2_+*ϵ*, or for notation convenience, as

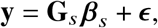

where we denote 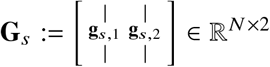, and 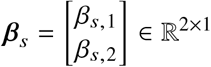 (similar for **G**_***t***_, *β*_***t***_). We can estimate the effect sizes of the tagging variant *t*,

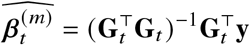

using the model **y** = **G**_***t***_ *β*_***t***_ + *ϵ*, and the expectation and variance of the estimated effects at variant *t* are

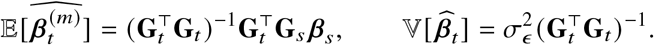

The derivation can be extended to multiple causal variants by replacing 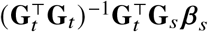 with 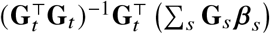, where the summation of *s* is over all causal variants.

To simplify the discussion, we further assume effects are the same across ancestries at causal variant *s, β* _*s*,1_ =*β* _*s*,2_ =*β*, and **g**_*s*_ := **g**_*s*,1_ + **g**_*s*,2_. Therefore,

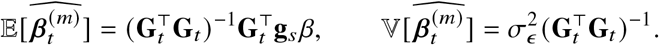

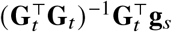 determines the expectation of the estimated effects at tagging variant *t*, and the differences in ancestry-specific taggability. Of note, 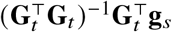 is exactly the solution of least squares when regressing **g**_*s*_ against **G**_*t*_; and if 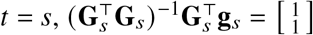 (which can be verified by noting **g**_*s*,1_ + **g**_*s*,2_ = **g**_*s*_).

#### Marginal effects-based methods for estimating heterogeneity

We describe details of marginal effects-based methods we used to estimate heterogeneity with input from a set of estimated effect sizes 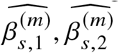 and corresponding estimated standard errors 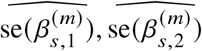 for a set of SNPs.

- Pearson correlation: obtained by calculating the Pearson correlation of 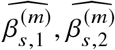 across SNPs. Pearson correlation does not model errors in estimated effects, therefore is expected be smaller than 1 and decreases with increasing magnitude of errors.
- OLS regression slope: obtained with OLS regression either by regressing 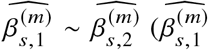 as the dependent variable,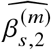 as the independent variable) or 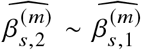. Similar to Pearson correlation, it does not model error terms in the independent variable. Moreover, it assumes homogeneous error terms in the dependent variable across observations. OLS regression slopes are susceptible to these errors and notably the results would vary when one exchange the regression orders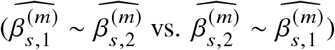 when 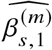 and 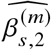 are associated with different standard errors ^46^ (as in the case for estimated effects in an admixed population with differential GWAS sample sizes).
- Deming regression slope: obtained with Deming regression ^34^ of 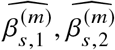, together with the estimated standard errors 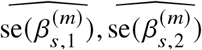. Deming regression models heterogeneous error terms in both the independent and dependent variables, therefore is more robust than Pearson correlation and OLS regression. Specifically, given a set of data and estimated standard errors (*x*_*i*_, *y*_*i*_, *σ*_*x,i*_, *σ*_*y,i*_), *i* = 1, …, *n* (we use a different set of notations for simplicity), Deming regression optimizes the following objective function to obtain estimated intercept *α* and slope *β*:

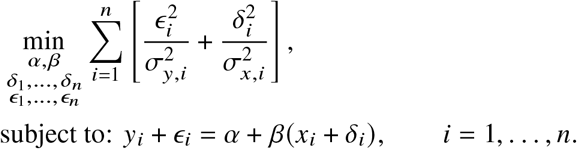

Standard errors of *α, β* can be obtained with bootstrapping. Of note, Deming regression slope will produce symmetric results with different regression orders (the obtained *β* slope will be reciprocal to each other). However, Deming regression is known to produce biased results when the standard errors *σ*_*x,i*_, *σ*_*y,i*_ are mis-specified ^46^.
- False positive rate of the HET test, as described above in “Parameter estimation and hypothesis testing”. It is expected to be well calibrated under the null, because its derivation as a likelihood ratio test. Similar to Deming regression, HET test properly models heterogeneous standard errors.

### Genotype data processing

#### PAGE genotype

We restricted our analysis within 17,299 genotyped individuals self-identified as African American in PAGE study ^1^. These individuals were from 3 studies: Women’s Health Initiative (WHI) (*n*=6,820), Multiethnic Cohort (MEC) (*n*=5,325) and the Icahn School of Medicine at Mount Sinai BioMe biobank in New York City (BioMe) (*n*=5,154). These individuals were genotyped on the Multi-Ethnic Genotyping Array (MEGA) which are designed to capture global genetic variation. More details on PAGE study can be found in previous publication ^1^. The genotype were imputed to the TOPMed reference panel and we performed standard quality control with imputation *R*^2^ *>* 0.8 and MAF *>* 0.5% to retain well-imputed variants. We calculated ancestry-specific (i.e. European and African ancestry-specific) allele frequencies within admixed individuals (see above) and further retained variants with MAF > 0.5% in both ancestries. This resulted in ∼6.9M variants and 17,299 individuals in our analysis.

#### UK Biobank genotype

We restricted our analysis within individuals with African-European admixed ancestries in UK Biobank. We first inferred the proportion of ancestries for each individual in UK Biobank using SCOPE ^47^ supervised using 1000 Genomes Phase 3 allele frequencies (AFR, EUR, EAS, SAS). We then selected African-European admixed individuals based on the inferred ancestry proportions. We retained 4,327 individuals with more than 5% of both AFR and EUR ancestries, and with less than 5% of both EAS and SAS ancestries. We filtered SNPs with imputation *R*^2^ *>* 0.8 and MAF > 0.5% to retain well-imputed variants. We retained variants with MAF > 0.5% in both ancestries. This resulted in ∼6.6M variants and 4,327 individuals in our analysis.

#### AoU genotype

We restricted our analysis within individuals with African-European admixed ancestries in AoU. First, we performed a principal component analysis of all 165,208 individuals in AoU microarray data (release v5) joint with 1,000 Genomes Phase 3 reference panel. Then we identified 31,375 individuals with African-European admixed ancestries (with at least both 10% European ancestries and 10% African ancestries, and who was within 2×normalized distance from the line connecting individuals of European ancestries and African ancestries in 1,000 Genomes reference panel) (Supplementary Notes). We performed basic quality control on genotypes of the identified individuals with African-European ancestries using PLINK2 with `--geno 0.05 --max-alleles 2 --maf 0.001`, and performed statistical phasing using Eagle2^48^ (v2.4.1) with default settings. We retained variants with MAF > 0.5% in both ancestries. This resulted in ∼0.65M variants and 31,375 individuals in our analysis. For AoU, we chose to use microarray data instead of whole genome sequencing data because microarray data of AoU contained more individuals and analyzing microarray data reduced the computational cost.

#### Local ancestry inference

We performed local ancestry inference using RFmix ^49^ (https://github.com/slowkoni/rfmix) with default parameters (8 generations since admixture). We used 99 CEU individuals and 108 YRI individuals from the 2,504 unrelated individuals in 1,000 Genome Project Phase 3^50^ as our reference populations. We used HapMap3 SNPs ^32^ when performing the local ancestry inference, and then interpolated the inferred local ancestry results to other variants in both PAGE and UK Biobank data sets. The accuracy of RFmix for local ancestry inference has been validated for African-European admixed individuals ^19^ (e.g., 98% accuracy for simulations with a realistic demographic model for African American individuals). We used the inferred local ancestry for both simulation study and real data analysis described below.

### Simulation study

We describe methods for simulation study that corresponds to each section of the results.

#### Pitfalls of including local ancestry in estimating heterogeneity

We first describe strategies of including local ancestry in estimating heterogeneity.

- For “lanc included”, we follow common practices ^17,19,51,52^ to use a term for local ancestry in Equation (1) *ℓ*_*s*_ (allele counts in African ancestries; defined above) as follows (restricting to SNP *s*):

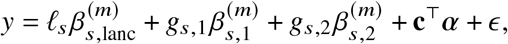

where 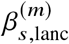 denotes the effect of local ancestry.
- For “lanc regressed”, we start with the equation 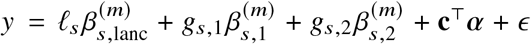 and we first estimate 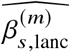 in the regression of 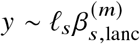, and then estimate 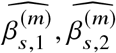 in regression of 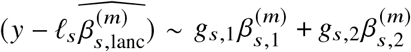.

To assess the impact of including local ancestry term when applying HET test to 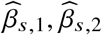 we randomly selected 1,000 SNPs on chromosome 1 in genotype with 17,299 PAGE individuals. We simulated traits with single causal variant. For each SNP, we simulated quantitative trait with the given SNP as the single causal variant with varying *β*_eur_ : *β*_afr_ = 1.0, 1.05, 1.1, 1.15, 1.2. We scaled *β*_eur_, *β*_afr_ such that the causal SNP explained the given amount of 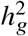. For each SNP, simulations of *β*_eur_, *β*_afr_ and environmental noises were repeated 30 times. We then applied different strategies of including local ancestry to these simulations and obtained *p*-value of HET testing *H*_0_ : *β*_eur_ =*β*_afr_. We additionally included the top principal component as a covariate throughout. We evaluated the distribution of false positive rate (FPR) or power of HET test by sub-sampling *without* replacement: we drew 100 random samples, each sample consisted of 500 SNPs, randomly drawn from the pool of 1,000 SNPs and 30 simulations; such sampling accounts for the randomness from both the environmental noises and SNP MAF. We calculated FPR or power for each sample of 500 SNPs, obtained empirical distributions of FPR or power (100 points each), and then calculated the mean and SE (using empirical standard deviation) from the empirical distribution.

#### Simulations with single causal variant

We performed simulations with single causal variant to assess the properties of methods based on estimated marginal effects. We randomly selected 100 regions each spanning 20 Mb on chromosome 1 (120K SNPs per region on average, SD 6K). For each region, the causal variant located at the middle of the region; it had same causal effects across local ancestries and was expected to explain a fixed amount of heritability (0.2%, 0.6%, 1.0%); the sign of the causal effect and environmental noises were randomly drawn 100 times. We evaluated the 4 metrics at both causal variants and clumped variants; clumped variants were obtained with regular LD clumping (index *p <* 5 ×10^−8^; *r*^2^ = 0.1, window size = 10 Mb) using PLINK: `--clump` function with parameters `--clump-p1 5e-8 --clump-p2 1e-4 --clump-r2 0.1--clump-kb 10000`. We used a 10Mb clumping window to account for the larger LD window within admixed individuals; other parameters were adopted from ref. ^53^. We found that when the simulated 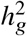 (of the single causal variant) was large, LD clumping can result in multiple SNPs because the secondary SNPs can reach *p <* 5×10^−8^ when we applied a commonly-used *r*^2^ = 0.1 threshold. Therefore, for each region, we either retained only the SNP with strongest association (matching the simulation setup of a single simulated causal variant), or retained all the SNPs from clumping results. Similar as above, we evaluated the distribution of 4 metrics by sub-sampling without replacement: we drew 100 random samples, each sample consisted of 500 regions (each region has 1 causal SNP), randomly drawn from the pool of 100 regions and 100 simulations; such sampling accounted for the randomness from both the environmental noises and SNP MAF. We then calculated the mean and SE from the 100 random samples.

#### Simulation with multiple causal variants

We performed simulations with multiple causal variants. We simulated multiple causal variants randomly distributed on chromosome 1 (515,087 SNPs). We drew *n*_causal_ = 62, 125, 250, 500, 1000 causal variants to simulate different level of polygenicity, such that on average there were approximately 0.25, 0.5, 1.0, 2.0, 4.0 causal variants per Mb. We fixed the heritability explained by all variants on chromosome 1 as 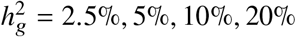. We performed sub-sampling without replacement to estimate the average and standard errors of the 4 metrics (each sample consisted of 1,000 SNPs, randomly drawn from SNPs across 500 simulations). We found that when the simulated 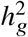 was small 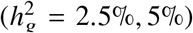, because the limited sample size in our data (*n* = 17, 299) for PAGE data, very few SNPs reach *p <* 5×10^−8^ in these simulations and consequently standard errors are very large and results can not be reliably reported. Therefore, we chose to report results only from 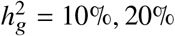 in Table S10.

#### Genome-wide simulation for evaluating our polygenic method

We performed simulations to evaluate our polygenic method in terms of parameter estimation of *r*_admix_ and hypothesis testing *H*_0_ : *r*_admix_ = 1 using real genome-wide genotypes. We simulated quantitative phenotypes using genotypes and inferred local ancestries with *N*=17,299 individuals and *S*=6,887,424 SNPs with MAF > 0.5% in both ancestries using the PAGE data set. The phenotypes were simulated under a wide range of genetic architectures varying proportion of causal variants *p*_causal_, heritability 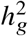, and true correlation *r*_admix_, and a frequency dependent effects distribution for causal variants: in each simulation, we first randomly drew *p*_causal_ proportion of causal variants. Given the set of causal variants, we simulated quantitative phenotypes based on Equations (3) and (4); 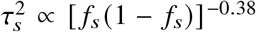 in Equation (4) where *α* = −0.38 obtained from a previous meta-analysis across 25 UK Biobank complex traits ^30^. The effect sizes were multiplied by a normalizing constant such that the variance explained by the genetic component 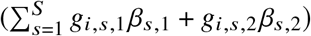 was equal to the desired heritability 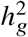. When estimating the genetic correlation, we either used all SNPs used in the simulation, or restricted to HapMap3 SNPs ^32^ to simulate scenarios where causal variants were not typed in the data. We applied our estimator as described in “Evaluation of genome-wide genetic effects consistency” using the same frequency dependent effects distribution used for phenotype simulation.

### Real data analysis

#### PAGE phenotype

We analyzed 24 heritable traits from PAGE study. The set of traits and diseases were the same set as analyzed in Extended Data Table 1 in ref. ^1^, except we did not separately analyze waist hip ratio for men and women. The only binary trait, type 2 diabetes, was modeled as a quantitative trait for convenience. We quantile normalized each trait, and included age, sex, age*sex, study center and top 10 in-sample principal components as covariates in the model. We also quantile normalized each covariate and used the average of each covariate to impute missing values in covariates.

#### UK Biobank phenotype

We analyzed 26 heritable traits from UKBB study. To select the set of traits to analyze, we first overlapped the UKBB traits we have access to with the set of traits analyzed in a previous paper ^54^ (that were selected based on heritability and proportion of individuals with non-missing phenotype values). Furthermore, we retained traits that have non-missing phenotype values for more than 1,000 individuals. After obtaining the 26 traits, for each trait, we quantile normalized phenotype values, and included age, sex, age*sex, and top 10 in-sample principal components as covariates in the model. We also quantile normalized each covariate and used the average of each covariate to imputed missing values in covariates.

#### AoU phenotype

We analyzed 10 heritable traits from AoU. The 10 traits included physical measurement and lipid phenotypes, which are straightforward to phenotype and have large sample sizes. Physical measurement phenotypes were extracted from Participant Provided Information in AoU dataset. Lipid phenotypes (including LDL, HDL, TC, TG) were extracted following github.com/all-of-us/ukb-cross-analysis-demo-project/tree/main/aou_workbench_siloed_analyses, including procedures of extracting most recent measurements per person, and correcting value with statin usage. After obtaining the 10 traits, for each trait, we quantile normalized phenotype values, and included age, sex, age*sex, and top 10 in-sample principal components as covariates in the model. We also quantile normalized each covariate and used the average of each covariate to imputed missing values in covariates.

### Genome-wide genetic correlation estimation

We calculated **K**_1_, **K**_2_ matrices in Equation (5) using either imputed SNPs and HapMap3 SNPs (for PAGE and UKBB), or microarray SNPs (for AoU), and using either frequency-dependent or GCTA heritability models by specifying 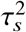. **K**_1_, **K**_2_ matrices were separately calculated for individuals within PAGE, UKBB, AoU studies. For each given *r*_admix_, we used GCTA software ^27^ to fit a single variance component model with the calculated **K**_1_+*r*_admix_**K**_2_ using `gcta64 --reml --reml-no-constrain`. We included age, sex, age*sex, top 10 in-sample principal components as covariates in analyzing all traits. We additionally included the causal signals at Duffy SNP (rs2814778) in 1q23.2 as covariates for analysis of white blood cell count and C-reactive protein because of the known strong admixture peak ^55,56^. Specifically, we used the local ancestries of SNP closest to Duffy SNP in our data as proxies for Duffy SNP (Duffy SNP itself is not typed or imputed in our data). The local ancestries are valid proxies of Duffy SNP because Duffy SNP is known to be highly differentiated across ancestries (alternate allele frequency is 0.006 v.s. 0.964 in ^50^) and therefore local ancestries are highly correlated with the Duffy SNP. We excluded closely related individuals in the analysis (< third-degree relatives; using ref. ^57^ with `plink2 --king-cutoff 0.0884`). In our main analyses, we used SNPs with MAF more than 0.5% in both ancestries (∼7M well-imputed SNPs for PAGE and UKBB; ∼0.65M array SNPs for AoU), and we assumed a frequency dependent effects distribution for causal variants. For PAGE and UKBB, we also performed secondary analyses with a smaller set of HM3 SNPs, and using other assumptions of effects distribution. We note that our meta-analysis credible interval across traits can be anti-conservative (i.e. the actual coverage probability is less than the nominal coverage probability) because we did not account for the genetic correlation across traits. We included the same covariates for the individual trait-SNP and statistical fine-mapping analyses described below.

### Individual trait-SNP analysis

We performed evaluation of genetic effects consistency at individual variants that were significantly associated with each trait. First, we performed GWAS for each study-trait pair with PLINK2^58^ `plink2 --linear --pheno-quantile-normalize --covar-quantile-normalize`. Second, we performed LD clumping with `plink --clump <assoc-file> --clump-p1 5e-8 -clump-p2 1e-4 -clump-r2 0.1 -clump-kb 10000`. For each clumped trait-SNP pair, we then obtained ancestry-specific estimated effect sizes and the corresponding standard errors.

### Statistical fine-mapping analysis

We performed fine-mapping analysis to each of the trait-SNP pair with significant heterogeneity by ancestry using SuSiE ^59^ (for PAGE and UKBB, for which we used genotype data with high SNP density). For each trait-SNP, we included all imputed SNPs in 3Mb window (1.5Mb upstream and downstream of the index SNP). We ran SuSiE with individual-level genotype and phenotype (covariates were regressed out of genotype and phenotype). We used default settings when running SuSiE: maximum number of 10 non-zero effects (`L = 10` in SuSiE parameter settings). And we obtained posterior inclusion probability and credible sets from the SuSiE output.

## Data Availability

All data produced in the present work are contained in the manuscript

## Data availability

PAGE individual-level genotype and phenotype data are available through dbGaP https://www.ncbi.nlm.nih.gov/projects/gap/cgi-bin/study.cgi?study_id=phs000356.v2.p1. UK Biobank individual-level genotype and phenotype data are available through application at http://www.ukbiobank.ac.uk. AoU individual-level genotype and phenotype are available through application at https://www.researchallofus.org.

## Code availability

Software implementing genome-wide genetic correlation estimation method: https://github.com/kangchenghou/admix-kit. Code for replicating analyses: https://github.com/kangchenghou/admix-genet-cor.

## Acknowledgements

We thank Alkes Price, Martin Jinye Zhang, Roshni Patel, Jonathan Pritchard, Arun Durvasula, Jianwen Cai, Ella Petter for helpful suggestions. This research was funded in part by the National Institutes of Health under awards R01-HG009120, R01-MH115676. This research was conducted using the UKBB Resource under application 33297. We thank the participants of UKBB for making this work possible. MESA and the MESA SHARe projects are conducted and supported by the National Heart, Lung, and Blood Institute (NHLBI) in collaboration with MESA investigators. Support for MESA is provided by contracts 75N92020D00001, HHSN268201500003I, N01-HC-95159, 75N92020D00005, N01-HC-95160, 75N92020D00002, N01-HC-95161, 75N92020D00003, N01-HC-95162, 75N92020D00006, N01-HC-95163, 75N92020D00004, N01-HC-95164, 75N92020D00007, N01-HC-95165, N01-HC-95166, N01-HC-95167, N01-HC-95168, N01-HC-95169, UL1-TR-000040, UL1-TR-001079, and UL1-TR-001420, UL1TR001881, and DK063491. Funding for SHARe genotyping was provided by NHLBI Contract N02-HL-64278. Genotyping was performed at Affymetrix (Santa Clara, California, USA) and the Broad Institute of Harvard and MIT (Boston, Massachusetts, USA) using the Affymetrix Genome-Wide Human SNP Array 6.0. The authors thank the other investigators, the staff, and the participants of the MESA study for their valuable contributions. A fill list of participating MESA investigators and institutes can be found at http://www.mesa-nhlbi.org. The Atherosclerosis Risk in Communities study has been funded in whole or in part with Federal funds from the National Heart, Lung, and Blood Institute, National Institutes of Health, Department of Health and Human Services, under Contract nos. (75N92022D00001, 75N92022D00002, 75N92022D00003, 75N92022D00004, 75N92022D00005). The authors thank the staff and participants of the ARIC study for their important contributions. The *All of Us* Research Program is supported by the National Institutes of Health, Office of the Director: Regional Medical Centers: 1 OT2 OD026549; 1 OT2 OD026554; 1 OT2 OD026557; 1 OT2 OD026556; 1 OT2 OD026550; 1 OT2 OD 026552; 1 OT2 OD026553; 1 OT2 OD026548; 1 OT2 OD026551; 1 OT2 OD026555; IAA #: AOD 16037; Federally Qualified Health Centers: HHSN 263201600085U; Data and Research Center: 5 U2C OD023196; Biobank: 1 U24 OD023121; The Participant Center: U24 OD023176; Participant Technology Systems Center: 1 U24 OD023163; Communications and Engagement: 3 OT2 OD023205; 3 OT2 OD023206; and Community Partners: 1 OT2 OD025277; 3 OT2 OD025315; 1 OT2 OD025337; 1 OT2 OD025276. In addition, the *All of Us* Research Program would not be possible without the partnership of its participants.

## Supplementary Information

Causal effects on complex traits are similar across segments of different continental ancestries within admixed individuals

### A Supplementary Notes

#### A.1 Heterogeneity by local ancestry in marginal effects in real data

Across 60 study-trait pairs, we detected a total of 217 GWAS significant clumped trait-SNP pairs and we estimated the ancestry-specific marginal effects for each of these SNPs at GWAS loci. 41 out of 217 trait-SNP pairs had significant heterogeneity in marginal effects by ancestry (HET test *p*_HET_ *<* 0.05 / 217). 16 out of 41 SNPs with significant heterogeneity were from UKBB data and PAGE data: 14 MCH-associated SNPs at 16p13.3 in UKBB data had strongest heterogeneity with average − log_10_ (*p*_HET_) = 13.0. By performing statistical fine-mapping analyses (Methods), we determined there were multiple conditionally independent association signals (Figure S3; also reported in ref. 36). Similarly, we determined there were multiple conditionally independent trait-associated variants nearby 1 RBC count-associated SNP at 16p13.3 in UKBB data (Figure S4; − log_10_ (*p*_HET_) = 5.5) and 1 CRP-associated SNPs at 1q23.2 in PAGE data (Figure S5; − log_10_ (*p*_HET_) = 4.1; also reported in ref. 56) that exhibited heterogeneity in marginal effects. The rest 25 out of 41 SNPs with significant heterogeneity were from AoU data: 22 height-associated SNPs with − log_10_ (*p*_HET_) = 6.8, 2 total cholesterol-associated SNPs with − log_10_ (*p*_HET_) = 5.8, 1 LDL-associated SNPs with average log_10_ − (*p*_HET_) = 6.8. We did not perform statistical fine-mapping on AoU microarray data, because we were concerned that imperfect tagging of relatively low density of microarray SNPs may lead to error-prone inference for existence of multiple causal variants. We leave fine-mapping analysis on high density SNPs (such as whole genome sequencing or densely-imputed) of AoU data for future work. Overall, we detected abundant evidence of multiple causal SNPs for loci that exhibit heterogeneity in marginal effects (especially for MCH-associated SNPs with the strongest heterogeneity), which was consistent to our simulation study with *r*_admix_ = 1 (see Results).

#### A.2 Local ancestry adjustment in heterogeneity estimation

We discuss the use of local ancestry in the heterogeneity estimation. Recall that our main equation is

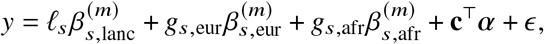

and we evaluated three approaches: (1) ignoring local ancestry altogether (“w/o”); (2) including local ancestry as a covariate in the model (“lanc-included”); (3) regressing out the local ancestry from phenotype (“lanc-regressed”) followed by heterogeneity estimation on residuals. In null simulations, we have observed the inflation of HET test using “lanc-regressed”. In power simulations, we have observed the reduced power of HET test using “lanc-included”.

These results are explained by the induced correlation between the local ancestry and ancestry-specific genotypes *𝓁*_*s*_, *g*_*s*,eur_, *g*_*s*,afr_. Intuitively, each additional local ancestry from African ancestries *𝓁*_*s*_, indicates an expected increase of risk allele counts from African ancestries *g*_*s*,afr_, and an expected decrease of risk allele counts from European ancestries *g*_*s*,eur_. Consequently, *𝓁*_*s*_ will be positively correlated with *g*_*s*,afr_ and negatively correlated with *g*_*s*,eur_. Indeed, the average correlation in a set of randomly sampled SNPs in PAGE data is 0.36 for *𝓁s* ∼ *g*_*s*,afr_ and #x2212; 0.55 for *𝓁s* ∼ *g*_*s*,eur_ (Table S12). Consequently, regressing out the local ancestry only from the phenotype is equivalent to adding a positive effect to *g*_*s*,eur_ and a negative effect to *g*_*s*,afr_; “lanc-regressed” leads to drastically inflated HET test (Figure 5a). On the other hand, a joint inference of *β*_*s*,lanc_, *β*_*s*,eur_, *β*_*s*,afr_ in the presence of correlations among *𝓁*_*s*_, *g*_*s*,eur_, *g*_*s*,afr_ would lead to increased variance in the estimated effects, therefore a power loss in “lanc-included” (Figure 5b).

#### A.3 Pitfalls of using marginal effects at GWAS significant variants to estimate heterogeneity

We investigated 4 methods that use marginal effects as input: (1) HET test; (2) Deming regression slopes of the marginal effects across SNPs (Deming slope); (3) Ordinary least squares regression slopes of estimated ancestry-specific marginal effects across SNPs (OLS slope); (4) Pearson correlation of the marginal effects across SNPs (Pearson correlation). Except for HET test which can compare effects difference for each individual SNP, the other 3 methods evaluate the aggregated effects difference across multiple SNPs. We have performed simulations both with single causal variant and multiple causal variants. In the following, we describe more details on the performance of HET test and Deming slope (in addition to those in Results section). And we also describe the performance of OLS slope and Pearson correlation.

##### Simulation with single causal variant

When evaluated at causal variants, in contrast to HET test and Deming slope, Pearson correlation and OLS slope were severely mis-calibrated (Figure S6 and table S9). For example, when the simulated 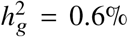, the false positive rate of HET test was 0.051 (SE 0.01), consistent with the expected level of 0.05 (Figure S6a); the average Deming regression slope was 1.005 (SE 0.01) when regressing *β*_eur_ against *β*afr (*β*_eur_ ∼*β*afr) and 0.996 (SE 0.01) for *β*_;afr;_ ∼ *β*_eur_, consistent with the expected slope of 1 (Figure S6bc). Pearson correlation significantly deviated from the expected level of 1: 0.964 (SE 0.005) (Figure S6); OLS slope was consistently smaller than 1 for *β*_afr_ ∼ *β*_eur_ (0.943 (SE 0.017)) and for *β*_eur_ ∼ *β*_afr_ (0.985 (SE 0.017)) (Figure S6ef). Interestingly, OLS slope’s downward bias varied with the regression order (*β*_afr_ ∼ *β*_eur_ v.s. *β*_eur_ ∼ *β*_afr;_). This is because the bias of OLS slope is a function of noise level in the independent variable, and estimated marginal effects in European ancestries and African ancestries were associated with different levels of standard errors (larger standard errors for *β*_eur_ because of smaller European ancestries proportion in PAGE African American individuals). Deming slope produced accurate results regardless of regression order as the differential standard errors are taken into consideration (Methods). Overall our results are consistent with mis-calibrations of Pearson correlation and OLS slopes due to their ignorance of the errors in the estimated effects 46.

When the causal SNPs were unknown and clumped SNP were used, as shown in Results section, HET test was increasingly mis-calibrated with larger 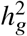 while Deming slope remained relatively robust (Figure 6). Similar to HET test, Pearson correlation and OLS slope were less calibrated for clumped SNPs, likely due to increased standard errors of estimated effects (Figure S6d-f). The mis-calibration induced by clumping arises from the inclusion of multiple SNPs in the clumped set (even though only 1 causal variant was simulated); the clumped SNPs included both index SNPs with the strongest associations, and secondary SNPs with weaker associations. These secondary SNPs were less correlated with the causal SNPs (average *r*^2^ = 0.072) and were physically more distant from the causal SNPs (average distance 432.5kb) compared to those strongest associated SNPs in each region (average *r*^2^ = 0.973, average distance 2.4kb) (in simulation with 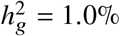), and therefore can induce heterogeneity by ancestry as indicated in Figure 1c. After restricting to SNPs with the strongest association after clumping (thus matching the simulation setup of a single simulated causal variant), both HET test and Deming slope resumed well-calibration (Table S9). This indicates the efficiency of LD clumping in capturing causal variant (e.g., 63% of clumped variants were causal when the simulated 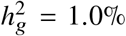; Table S11). However, we note OLS slope and Pearson correlation remained not calibrated (Table S9).

##### Simulation with multiple causal variants

In contrast to simulation with a single causal variant, even evaluated at the causal variants, both HET test and Deming slope were biased in the presence of multiple causals within the same LD region; the mis-calibration/bias increased with polygenicity (Figure 6 and table S10). For example, in simulation with 2 causal SNPs per Mb (*n*_causal_ = 500 on chromosome 1) and 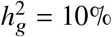, HET had inflated false positive rate (0.249 (SE 0.012) at the nominal 0.05 rate); average Deming slope of *β*_eur_ ∼ *β*_afr_ was 1.085 (SE 0.016). This is likely due to tagging among multiple causal variants whereby a causal SNP also tags effect of nearby causal SNPs in an ancestry-specific way (Methods). LD clumping did not alleviate the mis-calibration/bias (Figure 6 and table S10); for example, the average FPR was 0.279 (SE 0.008) and the average Deming slope was 1.083 (SE 0.018) in simulations with 4 causal variants per Mb (*n*_causal_ = 1, 000 on chromosome 1). Such mis-calibrations occurred irrespective of sample size (Figure S7), or simulated heritability 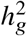 (Table S10). For completeness, we also evaluated OLS regression slope and Pearson correlation showing mis-calibrations with a large magnitude (Table S10). Finally, we note the upward/downward biases of Deming slope for *β*_eur_ ∼ *β*_afr_ / *β*_afr_ ∼ *β*_eur_, which were likely due to imbalanced ancestry proportions (∼80% African and ∼20% European ancestries) of admixed genotypes in PAGE data (Figure S8 and table S13).

#### A.4 Identifying individuals with admixed African and European ancestries with principal component analysis

We seek to identify individuals with African-European admixed ancestries from a diverse population of genotyped individuals (sample data, e.g., AoU) together with a reference panel (reference data, e.g., 1,000 Genomes) using principal component analysis. First, we perform a principal component analysis jointly on the sample and reference data and obtain top principal components (PCs) **w**_*i*_ for each individual *i*. We calculate the averaged PCs for individuals with European and African continental ancestries in 1,000 Genomes:

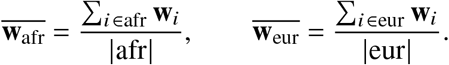

(In our analysis, for African continental ancestries in 1,000 Genomes, we did not include individuals with sup-population “ASW” (African Ancestry in Southwest US) or “ACB” (African Caribbean in Barbados), because some individuals in these sub-populations had admixed ancestries.) Second, we calculate the projected length and distance of each individual **w**_*i*_ to the line 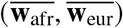. We first define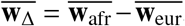, and calculate the normalized projected length *t*_*i*_ and distance *d*_*i*_ as:

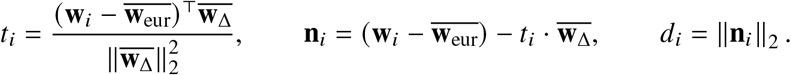

Roughly speaking, *t*_*i*_ ∈ [0, 1] if individual *i* locates in the PC space between European and African ancestries, and has the interpretation of global ancestry proportion: the closer *t*_*i*_ is to 1, the more African ancestries individual *i* has, and vice versa. Finally, we define the normalized distance 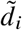 with

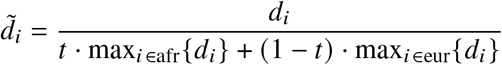

to account for the different spread (in PC space) for individuals with European and African continental ancestries. In our analysis, we used the first two PCs when calculating these quantities, and the set of selected admixed individuals was robust to the number of PCs used. We selected individuals with admixed ancestries with at least both 10% European ancestries and 10% African ancestries (*t*_*i*_∈ [0.1, 0.9]), and who was within 2 × normalized distance 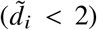 from the line connecting individuals of European ancestries and African ancestries in 1,000 Genomes reference panel.

### B Supplementary Figures

**Figure S1:**
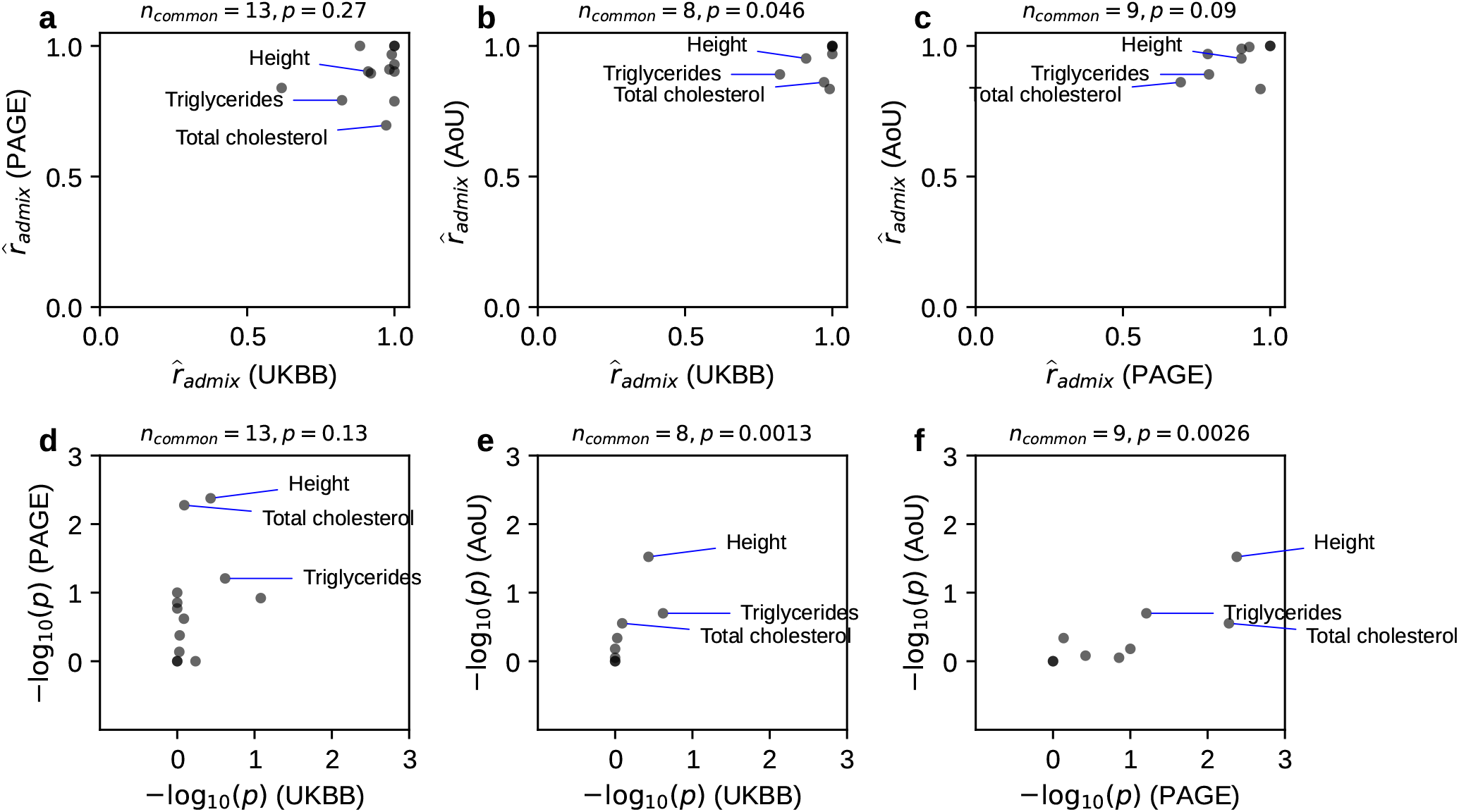
Consistency of *r*_admix_ across shared traits across studies. We compared estimated *r*_admix_ for shared traits across studies. We compared both 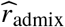 and − log_10_ (*p*) (for testing genetic correlation *H*_0_ : *r*_admix_ = 1). Three traits (Height, Triglycerides, Total cholesterol) with the most significant *p*-values for *H*_0_ : *r*_admix_ = 1 were annotated. Number of common traits shared across studies (*n*_common_) and Spearman correlation *p*-value were shown in the title for each panel. Overall, there were weak consistency of estimated 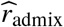 for shared traits across studies (although *p*-values for *H*_0_ : *r*_admix_ = 1 were consistent significantly). Numerical results are reported in Table S5.

**Figure S2:**
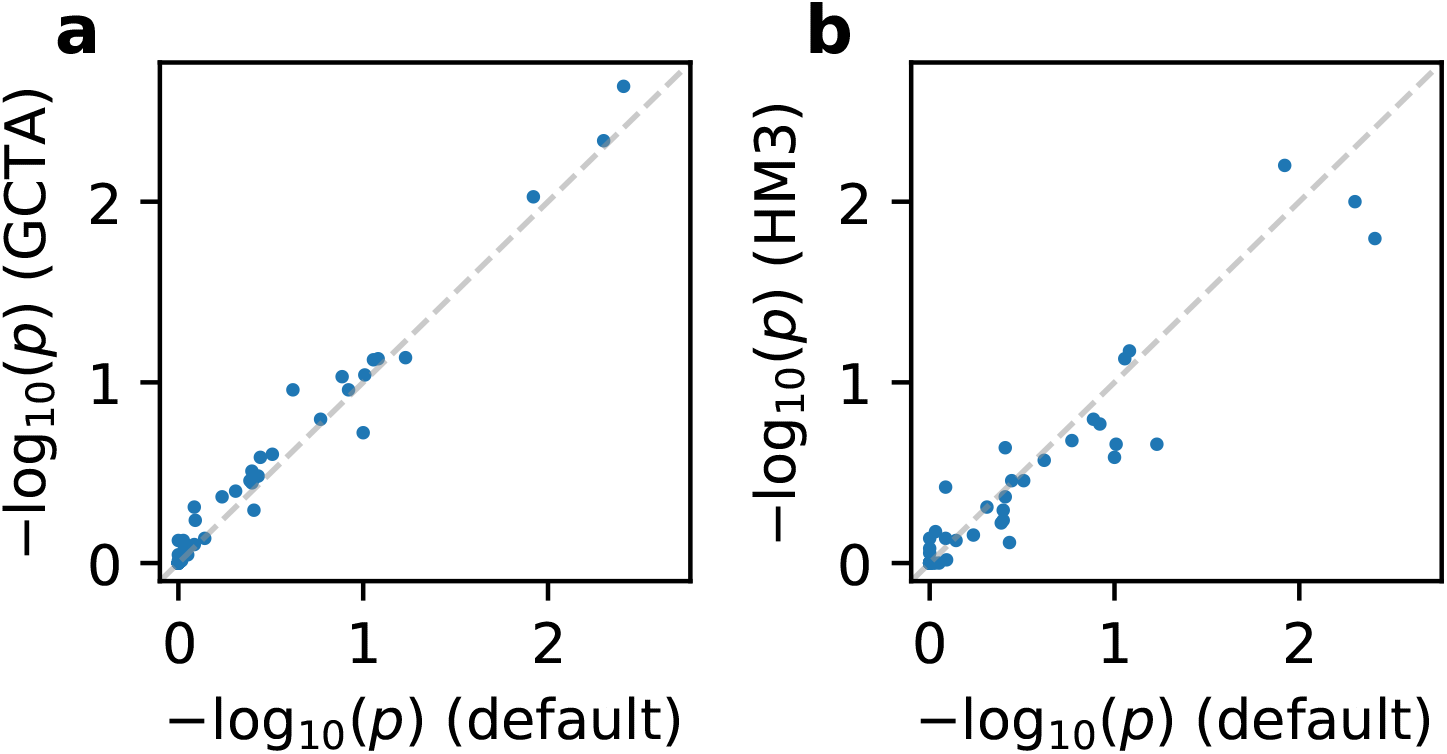
Genetic correlation estimation are robust to genetic architecture and SNP set. We performed *r*_admix_ estimation under the assumption of alternative genetic architecture and SNP set on real trait analysis across PAGE and UKBB. We compared *p*-values (for testing genetic correlation *H*_0_ : *r*_admix_ = 1) of our default setting (using frequency-dependent genetic architecture and imputed SNPs; Table 1) to those obtained using GCTA genetic architecture and imputed SNPs **(a)**, and to those obtained using frequency-dependent genetic architecture and HM3 SNPs **(b)**. Numerical results are reported in Table S6.

**Figure S3:**
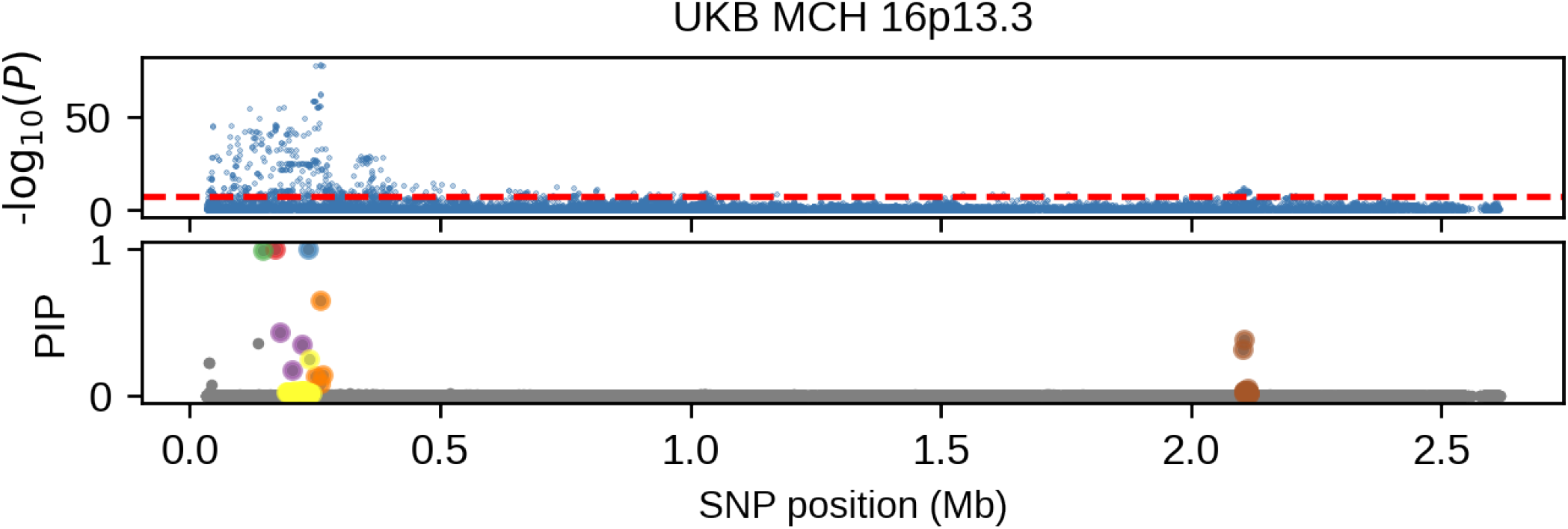
Association *p*-values and fine-mapping PIP of MCH at 16p13.3 for UK Biobank European-African admixed individuals. Upper panel corresponds to the association *p*-values and lower panel corresponds to the fine-mapping PIP. Different colors in the PIP plot corresponds to different credible sets.

**Figure S4:**
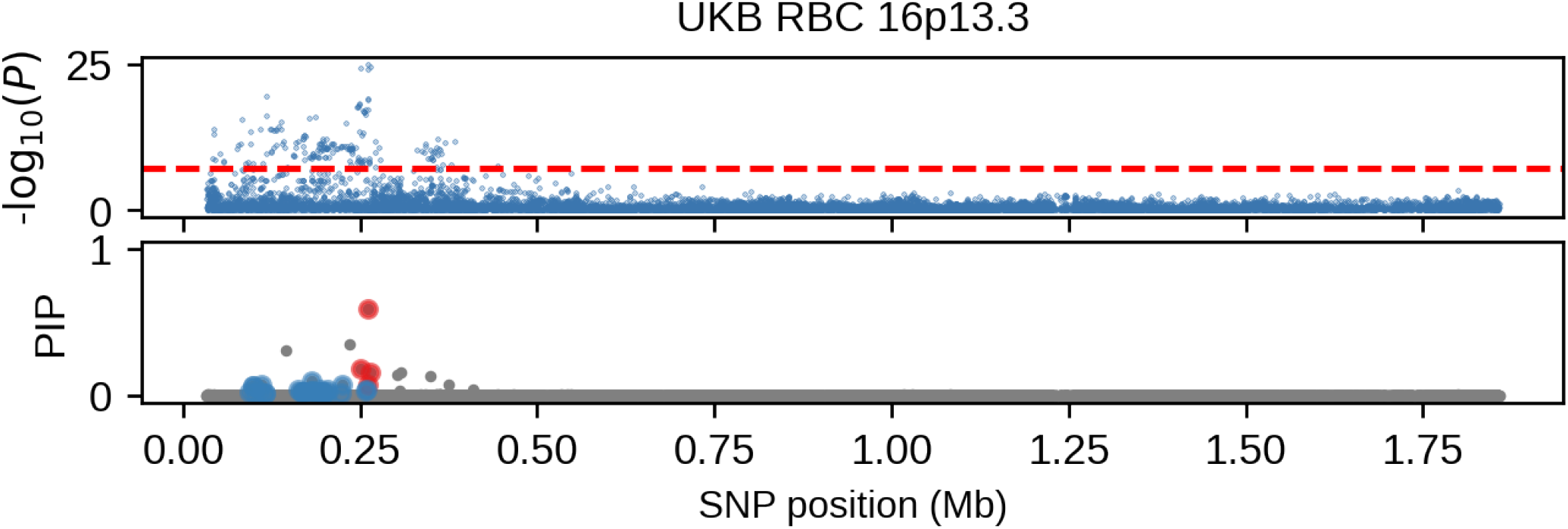
Association *p*-values and fine-mapping PIP of RBC at 16p13.3 for UK Biobank European-African admixed individuals. Upper panel corresponds to the association *p*-values and lower panel corresponds to the fine-mapping PIP. Different colors in the PIP plot corresponds to different credible sets.

**Figure S5:**
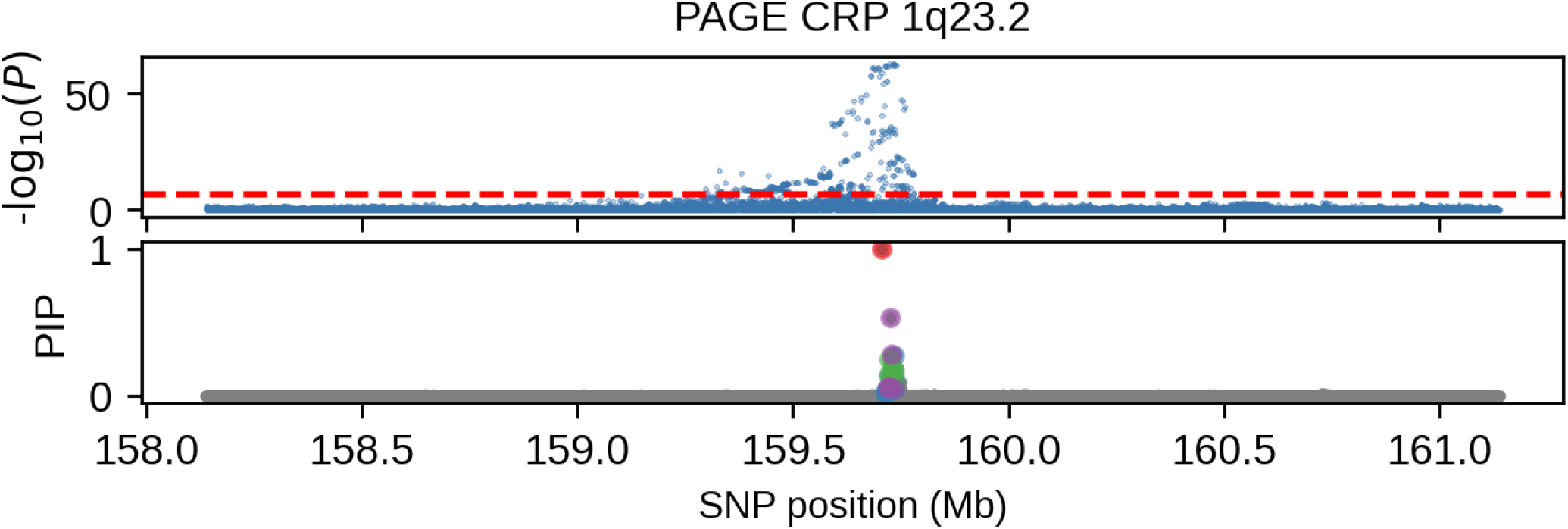
Association *p*-values and fine-mapping PIP of CRP at 1q23.2 for PAGE European-African admixed individuals. Upper panel corresponds to the association *p*-values and lower panel corresponds to the fine-mapping PIP. Different colors in the PIP plot corresponds to different credible sets.

**Figure S6:**
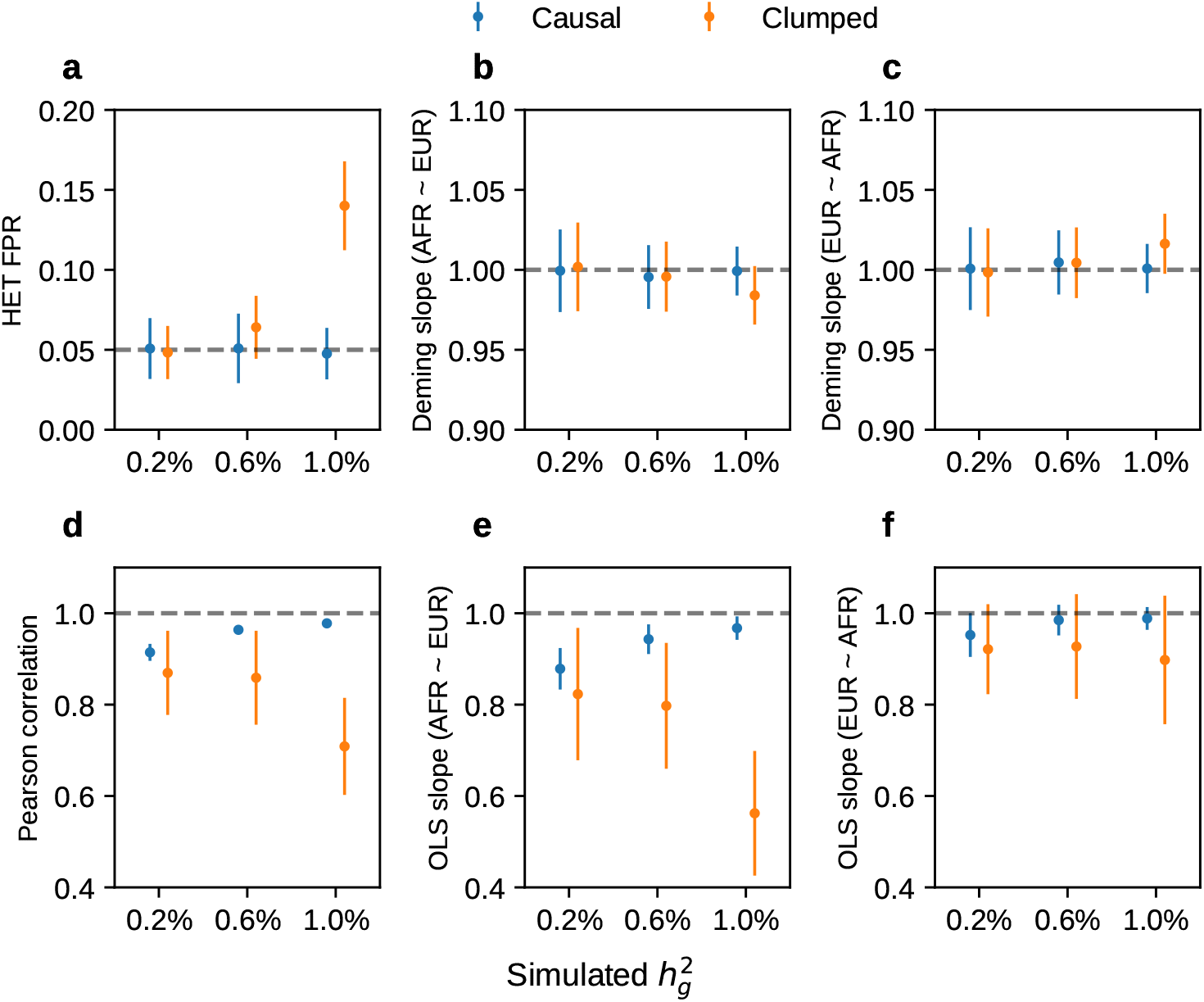
Simulations with single causal variant. Simulation were based on 100 regions each spanning 20 Mb on chromosome 1 and 17,299 PAGE individuals. In each simulation, we randomly selected single causal variant and simulated quantitative phenotypes where these causal variants had same causal effects across ancestries and each causal variant was expected to explain a fixed amount of heritability (0.2%, 0.6%, 1.0%). Each panel corresponds to one metric for both causal and clumped variants. **(a)** False positive rate (FPR) of HET test. **(b)** Deming regression slope with *β*_afr_ ∼ *β*_eur_. **(c)** Deming regression slope with *β*_eur_ ∼ *β*_afr_. **(d)** Pearson correlation. **(e)** OLS regression slope with *β*_afr_ ∼ *β*_eur_. **(f)** OLS regression slope with *β*_eur_ ∼ *β*_afr_. 95% confidence intervals were based on 100 random sub-samplings with each sample consisted of 500 SNPs (Methods). Numerical results are reported in Table S9.

**Figure S7:**
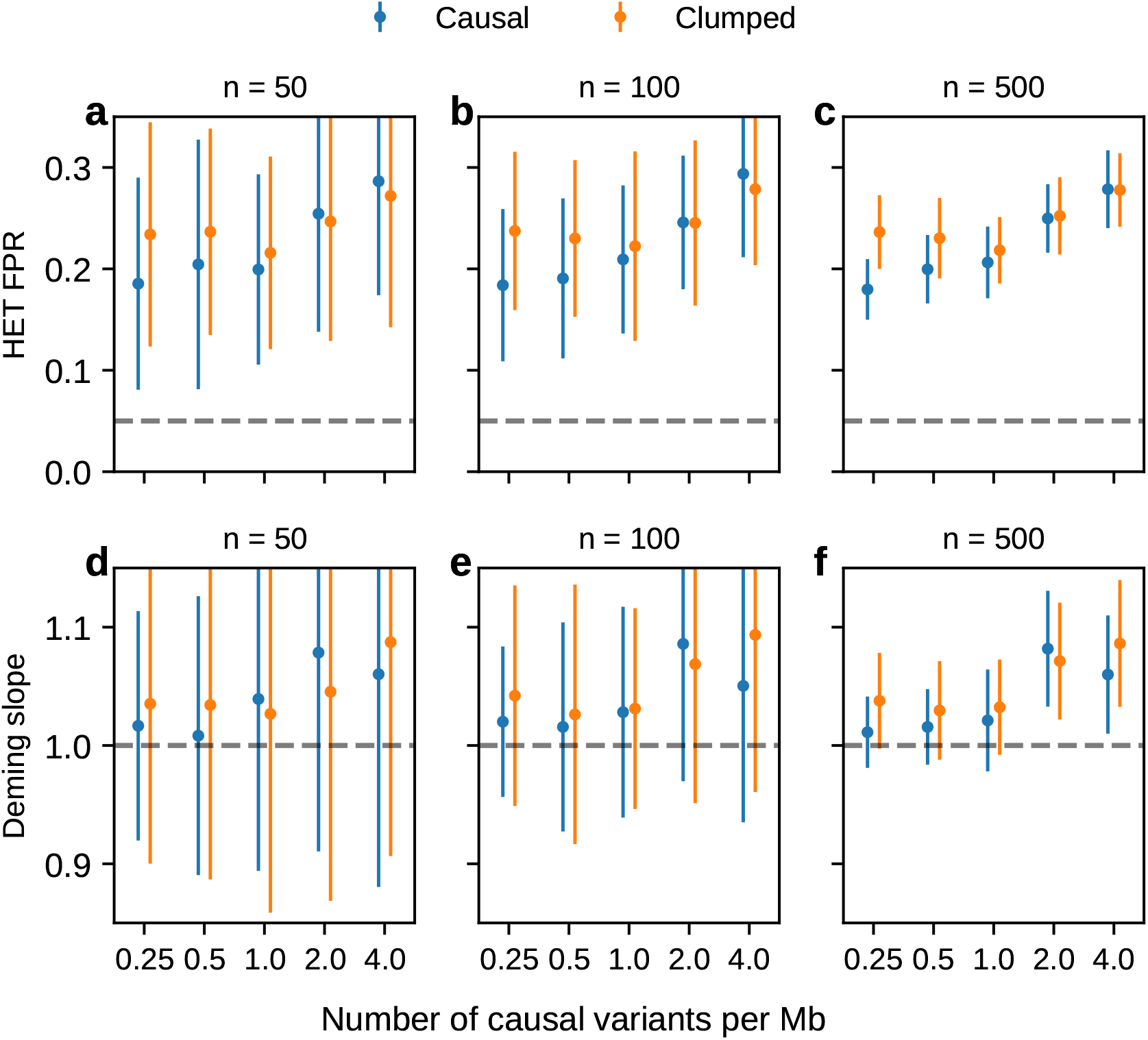
Simulation with multiple causal variants at other sample sizes (Figure 6cd). Simulation were based on chromosome 1 (515,087 SNPs) and 17,299 PAGE individuals. We drew 62, 125, 250, 500, 1000 causal variants to simulate different level of polygenicity, such that on average there were approximately 0.25, 0.5, 1.0, 2.0, 4.0 causal variants per Mb. The heritability explained by all causal variants was fixed at 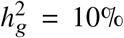. **(a-c)** False positive rate of HET test for the causal variants and clumped variants. **(d-f)** Deming regression slope of estimated ancestry-specific effects 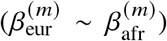 for the causal variants and clumped variants. 95% confidence intervals were based on 100 random sub-samplings with each sub-sample consisted of *n* = 50, 100, 500 SNPs (instead of *n* = 1, 000 SNPs in Figure 6cd) (Methods).

**Figure S8:**
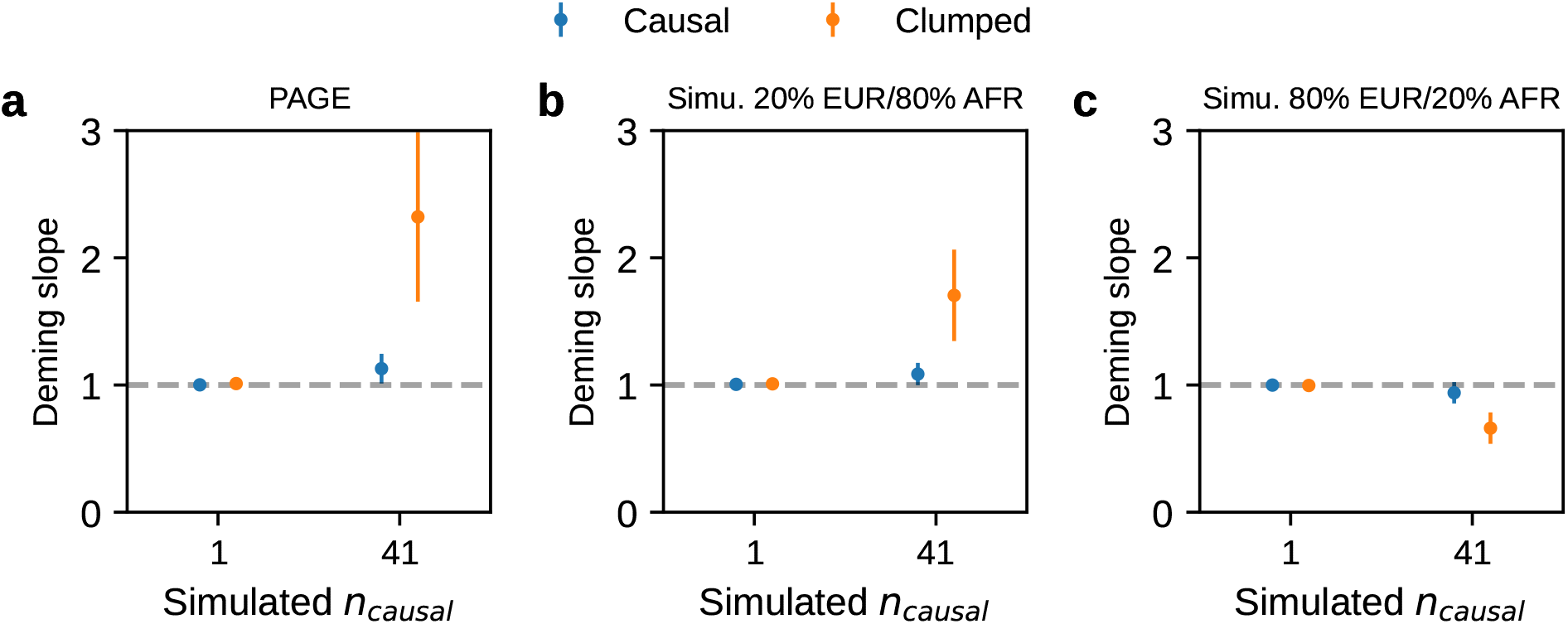
Simulation results in PAGE data set and simulated genotype data sets with varying ancestry proportion. We evaluated the bias of Deming slope 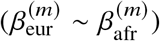 observed in Figure 6 by performing simulations on 3 genotype data sets: **(a)** PAGE data set of 17,299 individuals with ∼20% European and ∼80% African ancestry proportions. **(b)** simulated genotype data set of 20000 individuals with 20% European and 80% African ancestry proportions. **(c)** simulated genotype data set of 20,000 individuals with 80% European and 20% African ancestry proportions. Simulated genotype data was generated as follows: first, we simulated the local ancestries for each SNP and individual using a Poisson process parametrized by the recombination rate and genetic distance; second, we used the phased genotype segment from a random individual in 1,000 Genomes with the corresponding ancestry (European / African) as the genotype for each simulated local ancestry segment. Such simulation method preserves the realistic local ancestry segment length distribution, MAF and LD structure for the generated genotypes. To simulate the phenotype, we randomly selected 100 regions each spanning 20 Mb on chromosome 1. For each region, we either simulated *n*_causal_ = 1 causal variant at the middle of the region, or simulated *n*_causal_ = 41 equally spaced across 20 Mb (on average 2 causal variant per Mb); these causal variants had same causal effects across local ancestries and *each* causal variant was expected to explain a fixed amount of heritability (0.6%) (we simulated a large heritability to better simulate the bias due to different ancestry proportions). 95% confidence intervals were based on 100 random sub-samplings with each sample consisted of 500 SNPs. We determined that biases from PAGE and simulated genotype data with 20% European / 80% African ancestries were both upward and biases from simulated genotype data with 80% European / 20% African ancestries were downward. Therefore, we determined the biases were due to imbalanced ancestry proportions (∼ 20% European and ∼80% African ancestries) in PAGE data. Numerical results are reported in Table S13.

### C Supplementary Tables

**Table S1:**
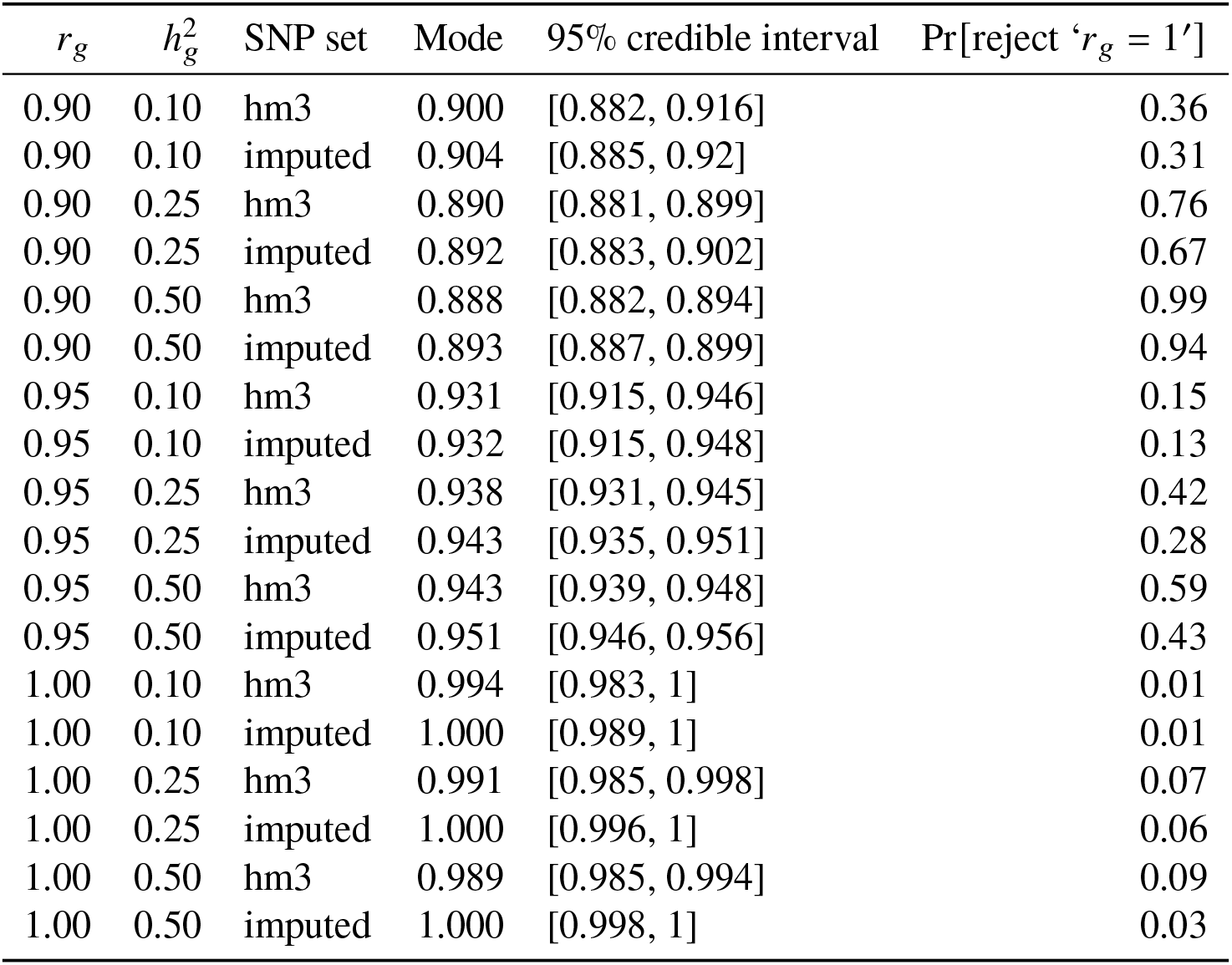
Numeric results of genetic correlation *r*_admix_ estimation in genome-wide simulations (with fixed *p*_causal_ = 0.1%; Figure 2). We fixed the proportion of causal variants *p*_causal_ = 0.1%, and we varied genome-wide heritability 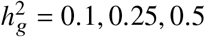, genetic correlation *r*_admix_ = 0.90, 0.95, 1.0, and SNP set used in the estimation. For each simulated genetic architecture, we performed a meta-analysis of estimation across 100 simulations. We report the mode and 95% credible interval from the meta-analysis. We also report the empirical probability of rejecting the null hypothesis of *r*_admix_ = 1.

**Table S2:** Numeric results of genetic correlation *r*_admix_ estimation in genome-wide simulations (with fixed 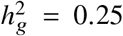). We fixed genome-wide heritability 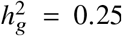, and we varied the proportion of causal variants *p*_causal_ = 0.001%, 0.01%, 0.1%, 1%, genetic correlation *r*_admix_ = 0.90, 0.95, 1.0, and SNP set used in the estimation. For each simulated genetic architecture, we performed a meta-analysis of estimation across 100 simulations, we report the mode and 95% credible interval from the meta-analysis. We also report the empirical probability of rejecting the null hypothesis of *r*_admix_ = 1.

| $r_g$ | $p_{\text{causal}}$ | SNP set | Mode | 95% credible interval | Pr[reject '$r_g = 1$'] |
| --- | --- | --- | --- | --- | --- |
| 0.90 | 0.001% | hm3 | 0.909 | [0.9, 0.917] | 0.61 |
| 0.90 | 0.001% | imputed | 0.911 | [0.901, 0.919] | 0.57 |
| 0.90 | 0.01% | hm3 | 0.900 | [0.891, 0.909] | 0.74 |
| 0.90 | 0.01% | imputed | 0.903 | [0.893, 0.912] | 0.59 |
| 0.90 | 0.1% | hm3 | 0.890 | [0.881, 0.899] | 0.76 |
| 0.90 | 0.1% | imputed | 0.892 | [0.883, 0.902] | 0.67 |
| 0.90 | 1% | hm3 | 0.902 | [0.893, 0.911] | 0.66 |
| 0.90 | 1% | imputed | 0.904 | [0.894, 0.913] | 0.59 |
| 0.95 | 0.001% | hm3 | 0.923 | [0.915, 0.931] | 0.52 |
| 0.95 | 0.001% | imputed | 0.930 | [0.921, 0.938] | 0.43 |
| 0.95 | 0.01% | hm3 | 0.944 | [0.937, 0.951] | 0.34 |
| 0.95 | 0.01% | imputed | 0.952 | [0.944, 0.959] | 0.31 |
| 0.95 | 0.1% | hm3 | 0.938 | [0.931, 0.945] | 0.42 |
| 0.95 | 0.1% | imputed | 0.943 | [0.935, 0.951] | 0.28 |
| 0.95 | 1% | hm3 | 0.939 | [0.932, 0.946] | 0.38 |
| 0.95 | 1% | imputed | 0.946 | [0.938, 0.954] | 0.28 |
| 1.00 | 0.001% | hm3 | 0.994 | [0.988, 0.999] | 0.16 |
| 1.00 | 0.001% | imputed | 1.000 | [0.996, 1] | 0.10 |
| 1.00 | 0.01% | hm3 | 0.988 | [0.982, 0.995] | 0.06 |
| 1.00 | 0.01% | imputed | 1.000 | [0.994, 1] | 0.03 |
| 1.00 | 0.1% | hm3 | 0.991 | [0.985, 0.998] | 0.07 |
| 1.00 | 0.1% | imputed | 1.000 | [0.996, 1] | 0.06 |
| 1.00 | 1% | hm3 | 0.994 | [0.989, 1] | 0.04 |
| 1.00 | 1% | imputed | 1.000 | [0.996, 1] | 0.02 |

**Table S3:** Numeric results of 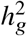 estimation in genome-wide simulations (with fixed *p*_causal_ = 0.1%). We fixed the proportion of causal variants *p*_causal_ = 0.1%, and we varied genome-wide heritability 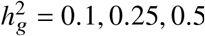, genetic correlation *r*_admix_ = 0.90, 0.95, 1.0, and SNP set used in the estimation. For each simulated genetic architecture, we report the mean and SEM of the estimates across 100 simulations.

| $h_g^2$ | $r_g$ | SNP set | Mean | SEM |
| --- | --- | --- | --- | --- |
| 0.10 | 0.90 | hm3 | 0.104 | 0.00218 |
| 0.10 | 0.90 | imputed | 0.11 | 0.0021 |
| 0.10 | 0.95 | hm3 | 0.0962 | 0.00203 |
| 0.10 | 0.95 | imputed | 0.101 | 0.00196 |
| 0.10 | 1.00 | hm3 | 0.0918 | 0.00204 |
| 0.10 | 1.00 | imputed | 0.0972 | 0.00197 |
| 0.25 | 0.90 | hm3 | 0.243 | 0.0025 |
| 0.25 | 0.90 | imputed | 0.259 | 0.0025 |
| 0.25 | 0.95 | hm3 | 0.239 | 0.00211 |
| 0.25 | 0.95 | imputed | 0.255 | 0.00219 |
| 0.25 | 1.00 | hm3 | 0.229 | 0.00199 |
| 0.25 | 1.00 | imputed | 0.247 | 0.00208 |
| 0.50 | 0.90 | hm3 | 0.468 | 0.0023 |
| 0.50 | 0.90 | imputed | 0.502 | 0.00223 |
| 0.50 | 0.95 | hm3 | 0.462 | 0.00221 |
| 0.50 | 0.95 | imputed | 0.498 | 0.00226 |
| 0.50 | 1.00 | hm3 | 0.46 | 0.0024 |
| 0.50 | 1.00 | imputed | 0.498 | 0.0024 |

**Table S4:** Numeric results of 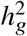 estimation in genome-wide simulations (with fixed 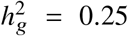). We fixed genome-wide heritability 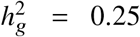, and we varied the proportion of causal variants *p*_causal_ = 0.001%, 0.01%, 0.1%, 1%, genetic correlation *r*_admix_ = 0.90, 0.95, 1.0, and SNP set used in the estimation. For each simulated genetic architecture, we report the mean and SEM of the estimates across 100 simulations.

| $p_{\text{causal}}$ | $r_g$ | SNP set | Mean | SEM |
| --- | --- | --- | --- | --- |
| 0.001% | 0.90 | hm3 | 0.24 | 0.0035 |
| 0.001% | 0.90 | imputed | 0.256 | 0.00377 |
| 0.001% | 0.95 | hm3 | 0.234 | 0.00374 |
| 0.001% | 0.95 | imputed | 0.252 | 0.00446 |
| 0.001% | 1.00 | hm3 | 0.235 | 0.00386 |
| 0.001% | 1.00 | imputed | 0.252 | 0.00448 |
| 0.01% | 0.90 | hm3 | 0.244 | 0.00219 |
| 0.01% | 0.90 | imputed | 0.261 | 0.00229 |
| 0.01% | 0.95 | hm3 | 0.239 | 0.00251 |
| 0.01% | 0.95 | imputed | 0.256 | 0.00252 |
| 0.01% | 1.00 | hm3 | 0.231 | 0.00246 |
| 0.01% | 1.00 | imputed | 0.249 | 0.00259 |
| 0.1% | 0.90 | hm3 | 0.243 | 0.0025 |
| 0.1% | 0.90 | imputed | 0.259 | 0.0025 |
| 0.1% | 0.95 | hm3 | 0.239 | 0.00211 |
| 0.1% | 0.95 | imputed | 0.255 | 0.00219 |
| 0.1% | 1.00 | hm3 | 0.229 | 0.00199 |
| 0.1% | 1.00 | imputed | 0.247 | 0.00208 |
| 1% | 0.90 | hm3 | 0.242 | 0.0023 |
| 1% | 0.90 | imputed | 0.257 | 0.0023 |
| 1% | 0.95 | hm3 | 0.238 | 0.00211 |
| 1% | 0.95 | imputed | 0.256 | 0.00211 |
| 1% | 1.00 | hm3 | 0.232 | 0.00204 |
| 1% | 1.00 | imputed | 0.248 | 0.00207 |

**Table S5:**
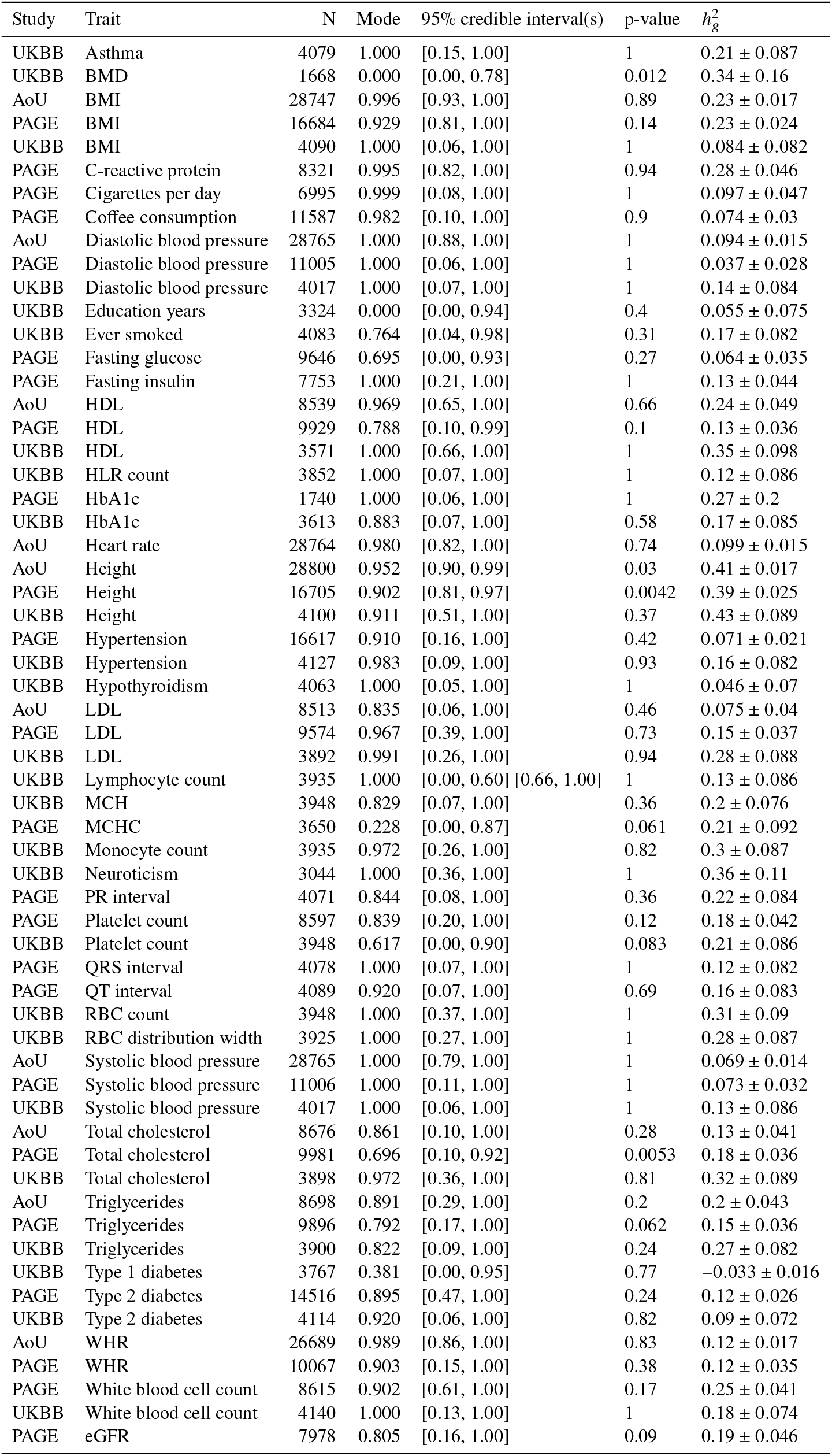
Genome-wide genetic correlation across 38 complex traits (60 study-trait pairs) for African-European admixed individuals in PAGE, UKBB, AoU. For each trait, we report number of individuals, posterior mode and 95% credible interval(s) for estimated *r*_admix_, *p*-value for rejecting the null hypothesis of *H*_0_ : *r*_admix_ = 1, and estimated heritability and standard error. Meta analysis results are performed across 60 study-trait pairs. UKBB Lymphocyte count has two credible intervals because of the non-concave profile likelihood curve, likely as a result of small sample size.

| Study | Trait | N | Mode | 95% credible interval(s) | p-value | $h_g^2$ |
| --- | --- | --- | --- | --- | --- | --- |
| UKBB | Asthma | 4079 | 1.000 | [0.15, 1.00] | 1 | 0.21 ± 0.087 |
| UKBB | BMD | 1668 | 0.000 | [0.00, 0.78] | 0.012 | 0.34 ± 0.16 |
| AoU | BMI | 28747 | 0.996 | [0.93, 1.00] | 0.89 | 0.23 ± 0.017 |
| PAGE | BMI | 16684 | 0.929 | [0.81, 1.00] | 0.14 | 0.23 ± 0.024 |
| UKBB | BMI | 4090 | 1.000 | [0.06, 1.00] | 1 | 0.084 ± 0.082 |
| PAGE | C-reactive protein | 8321 | 0.995 | [0.82, 1.00] | 0.94 | 0.28 ± 0.046 |
| PAGE | Cigarettes per day | 6995 | 0.999 | [0.08, 1.00] | 1 | 0.097 ± 0.047 |
| PAGE | Coffee consumption | 11587 | 0.982 | [0.10, 1.00] | 0.9 | 0.074 ± 0.03 |
| AoU | Diastolic blood pressure | 28765 | 1.000 | [0.88, 1.00] | 1 | 0.094 ± 0.015 |
| PAGE | Diastolic blood pressure | 11005 | 1.000 | [0.06, 1.00] | 1 | 0.037 ± 0.028 |
| UKBB | Diastolic blood pressure | 4017 | 1.000 | [0.07, 1.00] | 1 | 0.14 ± 0.084 |
| UKBB | Education years | 3324 | 0.000 | [0.00, 0.94] | 0.4 | 0.055 ± 0.075 |
| UKBB | Ever smoked | 4083 | 0.764 | [0.04, 0.98] | 0.31 | 0.17 ± 0.082 |
| PAGE | Fasting glucose | 9646 | 0.695 | [0.00, 0.93] | 0.27 | 0.064 ± 0.035 |
| PAGE | Fasting insulin | 7753 | 1.000 | [0.21, 1.00] | 1 | 0.13 ± 0.044 |
| AoU | HDL | 8539 | 0.969 | [0.65, 1.00] | 0.66 | 0.24 ± 0.049 |
| PAGE | HDL | 9929 | 0.788 | [0.10, 0.99] | 0.1 | 0.13 ± 0.036 |
| UKBB | HDL | 3571 | 1.000 | [0.66, 1.00] | 1 | 0.35 ± 0.098 |
| UKBB | HLR count | 3852 | 1.000 | [0.07, 1.00] | 1 | 0.12 ± 0.086 |
| PAGE | HbA1c | 1740 | 1.000 | [0.06, 1.00] | 1 | 0.27 ± 0.2 |
| UKBB | HbA1c | 3613 | 0.883 | [0.07, 1.00] | 0.58 | 0.17 ± 0.085 |
| AoU | Heart rate | 28764 | 0.980 | [0.82, 1.00] | 0.74 | 0.099 ± 0.015 |
| AoU | Height | 28800 | 0.952 | [0.90, 0.99] | 0.03 | 0.41 ± 0.017 |
| PAGE | Height | 16705 | 0.902 | [0.81, 0.97] | 0.0042 | 0.39 ± 0.025 |
| UKBB | Height | 4100 | 0.911 | [0.51, 1.00] | 0.37 | 0.43 ± 0.089 |
| PAGE | Hypertension | 16617 | 0.910 | [0.16, 1.00] | 0.42 | 0.071 ± 0.021 |
| UKBB | Hypertension | 4127 | 0.983 | [0.09, 1.00] | 0.93 | 0.16 ± 0.082 |
| UKBB | Hypothyroidism | 4063 | 1.000 | [0.05, 1.00] | 1 | 0.046 ± 0.07 |
| AoU | LDL | 8513 | 0.835 | [0.06, 1.00] | 0.46 | 0.075 ± 0.04 |
| PAGE | LDL | 9574 | 0.967 | [0.39, 1.00] | 0.73 | 0.15 ± 0.037 |
| UKBB | LDL | 3892 | 0.991 | [0.26, 1.00] | 0.94 | 0.28 ± 0.088 |
| UKBB | Lymphocyte count | 3935 | 1.000 | [0.00, 0.60] [0.66, 1.00] | 1 | 0.13 ± 0.086 |
| UKBB | MCH | 3948 | 0.829 | [0.07, 1.00] | 0.36 | 0.2 ± 0.076 |
| PAGE | MCHC | 3650 | 0.228 | [0.00, 0.87] | 0.061 | 0.21 ± 0.092 |
| UKBB | Monocyte count | 3935 | 0.972 | [0.26, 1.00] | 0.82 | 0.3 ± 0.087 |
| UKBB | Neutroicisim | 3044 | 1.000 | [0.36, 1.00] | 1 | 0.36 ± 0.11 |
| PAGE | PR interval | 4071 | 0.844 | [0.08, 1.00] | 0.36 | 0.22 ± 0.084 |
| PAGE | Platelet count | 8597 | 0.839 | [0.20, 1.00] | 0.12 | 0.18 ± 0.042 |
| UKBB | Platelet count | 3948 | 0.617 | [0.00, 0.90] | 0.083 | 0.21 ± 0.086 |
| PAGE | QRS interval | 4078 | 1.000 | [0.07, 1.00] | 1 | 0.12 ± 0.082 |
| PAGE | QT interval | 4089 | 0.920 | [0.07, 1.00] | 0.69 | 0.16 ± 0.083 |
| UKBB | RBC count | 3948 | 1.000 | [0.37, 1.00] | 1 | 0.31 ± 0.09 |
| UKBB | RBC distribution width | 3925 | 1.000 | [0.27, 1.00] | 1 | 0.28 ± 0.087 |
| AoU | Systolic blood pressure | 28765 | 1.000 | [0.79, 1.00] | 1 | 0.069 ± 0.014 |
| PAGE | Systolic blood pressure | 11006 | 1.000 | [0.11, 1.00] | 1 | 0.073 ± 0.032 |
| UKBB | Systolic blood pressure | 4017 | 1.000 | [0.06, 1.00] | 1 | 0.13 ± 0.086 |
| AoU | Total cholesterol | 8676 | 0.861 | [0.10, 1.00] | 0.28 | 0.13 ± 0.041 |
| PAGE | Total cholesterol | 9981 | 0.696 | [0.10, 0.92] | 0.0053 | 0.18 ± 0.036 |
| UKBB | Total cholesterol | 3898 | 0.972 | [0.36, 1.00] | 0.81 | 0.32 ± 0.089 |
| AoU | Triglycerides | 8698 | 0.891 | [0.29, 1.00] | 0.2 | 0.2 ± 0.043 |
| PAGE | Triglycerides | 9896 | 0.792 | [0.17, 1.00] | 0.062 | 0.15 ± 0.036 |
| UKBB | Triglycerides | 3900 | 0.822 | [0.09, 1.00] | 0.24 | 0.27 ± 0.082 |
| UKBB | Type 1 diabetes | 3767 | 0.381 | [0.00, 0.95] | 0.77 | -0.033 ± 0.016 |
| PAGE | Type 2 diabetes | 14516 | 0.895 | [0.47, 1.00] | 0.24 | 0.12 ± 0.026 |
| UKBB | Type 2 diabetes | 4114 | 0.920 | [0.06, 1.00] | 0.82 | 0.09 ± 0.072 |
| AoU | WHR | 26689 | 0.989 | [0.86, 1.00] | 0.83 | 0.12 ± 0.017 |
| PAGE | WHR | 10067 | 0.903 | [0.15, 1.00] | 0.38 | 0.12 ± 0.035 |
| PAGE | White blood cell count | 8615 | 0.902 | [0.61, 1.00] | 0.17 | 0.25 ± 0.041 |
| UKBB | White blood cell count | 4140 | 1.000 | [0.13, 1.00] | 1 | 0.18 ± 0.074 |
| PAGE | eGFR | 7978 | 0.805 | [0.16, 1.00] | 0.09 | 0.19 ± 0.046 |

<u>See Supplementary Excel table.</u>

Table S6: **Genetic correlation estimation are robust to genetic architecture and SNP set**. We performed *r*_admix_ estimation under the assumption of alternative genetic architecture and SNP set on real trait analysis across PAGE and UKBB. We compared posterior mode, 95% credible intervals and *p*-values (for testing *r*_admix_ *<* 1) of our default setting (using frequency-dependent genetic architecture and imputed SNPs; Table 1) to those obtained using GCTA genetic architecture and imputed SNPs, and to those obtained using frequency-dependent genetic architecture and HM3 SNPs. Study-trait pairs whose GCTA optimization failed to converge are indicated as ‘NA’.

<u>See Supplementary Excel table.</u>

Table S7: **Summary statistics of 217 genome-wide significant trait-associated SNPs**. We performed GWAS for each of 60 study-trait pairs in PAGE, UKBB, AoU. We report summary statistics for each of trait-associated SNPs that have association *p <* 5 × 10^−8^ and minor allele frequency > 0.5% in both European and African ancestries. For each pair of trait and SNP, we report association *p*-value, HET *p*-value, ancestry-specific allele frequencies calculated within PAGE, UKBB, AoU admixed individuals, estimated ancestry-specific effect sizes and standard errors. Across all 217 SNPs, Pearson’s *r* = 0.73 (SE 0.04), OLS regression slope of *β*_afr_ ∼ *β*_eur_ = 0.84 (SE 0.06), OLS regression slope of *β*_eur_ ∼ *β*_afr_ = 0.64 (SE 0.06), Deming slope of *β*_afr_ ∼ *β*_eur_ = 1.22 (SE 0.09), Deming regression slope of *β*_eur_ ∼ *β*_afr_ = 0.82 (SE 0.06). Across 193 SNPs after excluding MCH-associated SNPs, Pearson’s *r* = 0.85 (SE 0.03), OLS regression slope of *β*_afr_ ∼ *β*_eur_ = 0.92 (SE 0.05), OLS regression slope of *β*_eur_ ∼ *β*_afr_ = 0.80 (SE 0.06), Deming slope of *β*_afr_ ∼ *β*_eur_ = 1.08 (SE 0.05), Deming regression slope of *β*_eur_ ∼ *β*_afr_ = 0.93 (SE 0.04).

**Table S8:** Numerical results for pitfalls of including local ancestry in estimating heterogeneity (Figure 5). We report the false positive rate (for null simulations) and power (for power simulations) for HET *p*-values. In each simulation, we selected a single causal variant and simulated quantitive phenotypes where causal variants had varying heritability 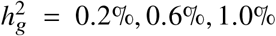 and varying ratios of effects across ancestries *β*_eur_ : *β*_afr_ = 1.0, 1.05, 1.1, 1.15, 1.2. Standard errors (displayed in parentheses) were calculated based on 100 random sub-samplings with each sample consisting of 500 SNPs (Methods).

| $h_g^2$ | $\beta_{\text{eur}} : \beta_{\text{afr}}$ | w/o | lanc included | lanc regressed |
| --- | --- | --- | --- | --- |
| 0.2% | 1.00 | 0.0515 (0.012) | 0.0509 (0.01) | 0.0768 (0.013) |
|  | 1.05 | 0.0525 (0.011) | 0.0514 (0.0096) | 0.0791 (0.014) |
|  | 1.10 | 0.0598 (0.01) | 0.0575 (0.0094) | 0.0814 (0.014) |
|  | 1.15 | 0.0714 (0.011) | 0.0613 (0.011) | 0.0884 (0.014) |
|  | 1.20 | 0.0826 (0.011) | 0.0675 (0.01) | 0.0946 (0.014) |
| 0.6% | 1.00 | 0.0503 (0.01) | 0.0509 (0.0094) | 0.166 (0.014) |
|  | 1.05 | 0.0583 (0.011) | 0.0557 (0.01) | 0.174 (0.016) |
|  | 1.10 | 0.0841 (0.012) | 0.0687 (0.013) | 0.191 (0.015) |
|  | 1.15 | 0.113 (0.015) | 0.0832 (0.013) | 0.207 (0.018) |
|  | 1.20 | 0.153 (0.016) | 0.104 (0.013) | 0.22 (0.019) |
| 1.0% | 1.00 | 0.0524 (0.01) | 0.0528 (0.0091) | 0.228 (0.018) |
|  | 1.05 | 0.0631 (0.01) | 0.0584 (0.0097) | 0.242 (0.018) |
|  | 1.10 | 0.0989 (0.013) | 0.0771 (0.012) | 0.263 (0.021) |
|  | 1.15 | 0.158 (0.015) | 0.106 (0.015) | 0.283 (0.022) |
|  | 1.20 | 0.227 (0.017) | 0.147 (0.017) | 0.308 (0.02) |

**Table S9:** Numerical results for simulations with single causal variant (Figure 6). We report the average and standard errors for each metric in simulations. Standard errors (displayed in parentheses) are based on 100 random sub-samplings with each sample consists of 500 SNPs. For clumped variants, we either retained only the SNP with strongest association within each region (“Clumped (single)”), or retained all the SNPs from clumping results (“Clumped (all)”) (Methods).

| group | $h_g^2$ | HET FPR | Deming (AFR~EUR) | Deming (EUR~AFR) | Pearson r | OLS (AFR~EUR) | OLS (EUR~AFR) |
| --- | --- | --- | --- | --- | --- | --- | --- |
| Causal | 0.2% | 0.051 (0.01) | 0.999 (0.013) | 1.001 (0.013) | 0.914 (0.009) | 0.878 (0.023) | 0.952 (0.024) |
|  | 0.6% | 0.051 (0.011) | 0.996 (0.01) | 1.005 (0.01) | 0.964 (0.005) | 0.943 (0.017) | 0.985 (0.017) |
|  | 1.0% | 0.048 (0.008) | 0.999 (0.008) | 1.001 (0.008) | 0.978 (0.002) | 0.967 (0.013) | 0.989 (0.013) |
| Clumped (single) | 0.2% | 0.048 (0.008) | 1.003 (0.014) | 0.997 (0.014) | 0.886 (0.048) | 0.837 (0.08) | 0.939 (0.032) |
|  | 0.6% | 0.049 (0.009) | 0.998 (0.009) | 1.002 (0.009) | 0.964 (0.004) | 0.947 (0.017) | 0.983 (0.017) |
|  | 1.0% | 0.047 (0.009) | 0.999 (0.007) | 1.001 (0.007) | 0.978 (0.002) | 0.968 (0.013) | 0.988 (0.013) |
| Clumped (all) | 0.2% | 0.048 (0.008) | 1.002 (0.014) | 0.998 (0.014) | 0.87 (0.047) | 0.823 (0.074) | 0.921 (0.05) |
|  | 0.6% | 0.064 (0.01) | 0.996 (0.011) | 1.004 (0.011) | 0.859 (0.052) | 0.797 (0.07) | 0.927 (0.058) |
|  | 1.0% | 0.14 (0.014) | 0.984 (0.009) | 1.016 (0.01) | 0.709 (0.054) | 0.562 (0.07) | 0.898 (0.072) |

**Table S10:**
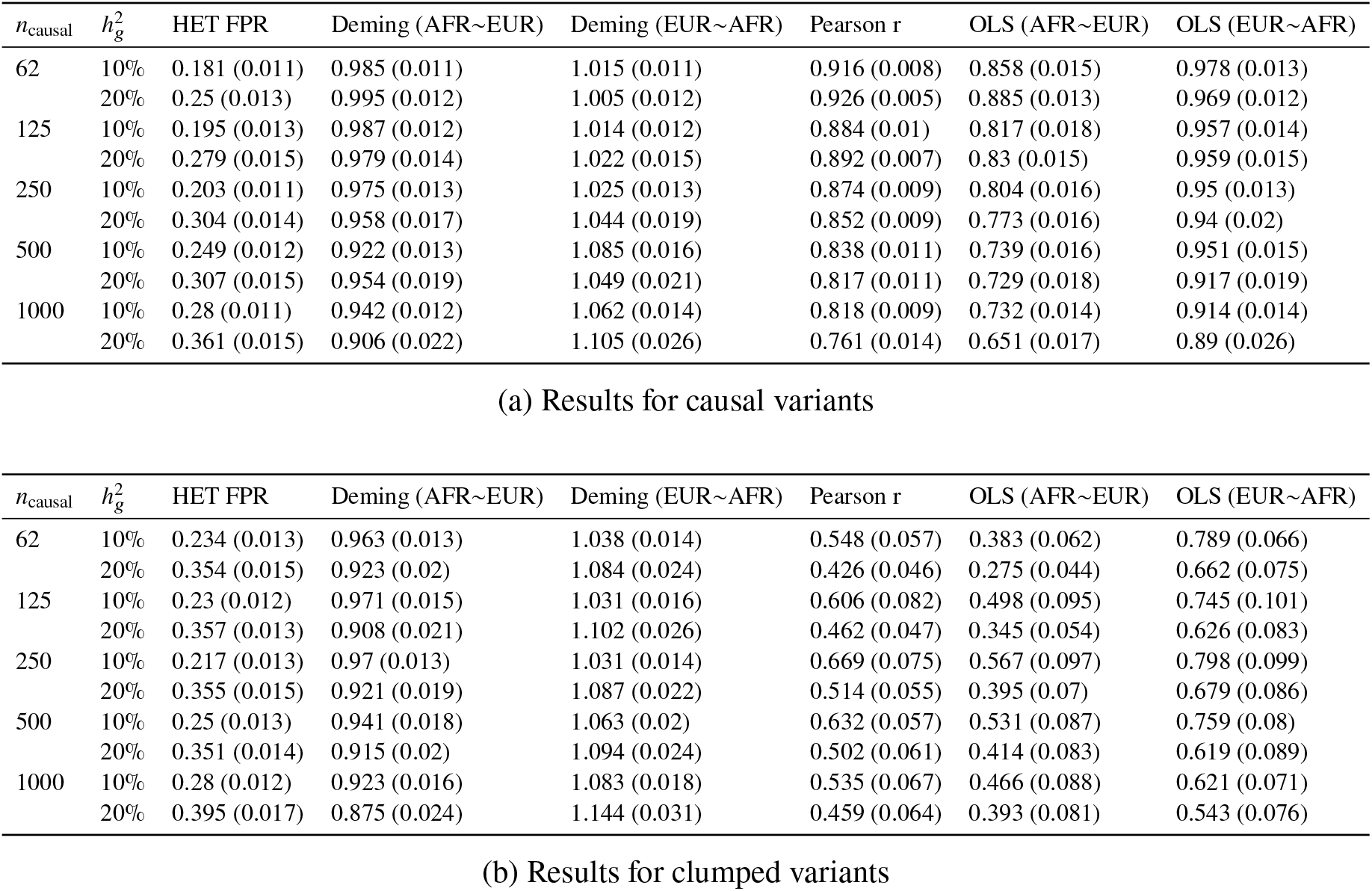
Numerical results for simulations with multiple causal variants (Figure 6). We report numerical results for the 4 metrics in simulations with varying number of causal variants 62, 125, 250, 500, 1000 causal variants (such that on average there were approximately 0.25, 0.5, 1.0, 2.0, 4.0 causal variants per Mb) and varying heritability explained by all causal variants 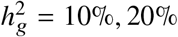. We specified heritability values that were larger than the chromosome 1 heritability of a typical complex trait because the limited sample size in our data produced only few clumped variants when 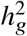 was small. We report the average and standard errors for each metric in simulations. Standard errors (displayed in parentheses) were based on 100 random sub-samplings with each sample consists of 1,000 SNPs (Methods).

**Table S11:** Probability for the clumped variants being causal in simulations with single causal variant (Figure 6). We report the probability for the clumped variants to be causal in simulations. “Single” corresponds to retaining only the SNP with strongest association within each region and “All” corresponds to retaining all the SNPs from clumping results (Methods).

| $h_g^2$ | Single | All |
| --- | --- | --- |
| 0.2% | 0.45 | 0.45 |
| 0.6% | 0.57 | 0.51 |
| 1.0% | 0.63 | 0.36 |

**Table S12:** Correlation between ancestry-specific genotypes and local ancestry. We report the pairwise Pearson correlation across ancestry-specific genotypes *g*_*s*,eur_, *g*_*s*,eur_ and local ancestry *ℓ*_*s*_. For each SNP, we calculated the pairwise correlations across 17,299 individuals. We report the mean and SEM averaged across 500 random SNPs across chromosome 1.

|  | EUR | AFR | lanc |
| --- | --- | --- | --- |
| EUR | 1.00 | -0.22 ± 0.0086 | -0.55 ± 0.0098 |
| AFR | -0.22 ± 0.0086 | 1.00 | 0.36 ± 0.0092 |
| lanc | -0.55 ± 0.0098 | 0.36 ± 0.0092 | 1.00 |

**Table S13:**
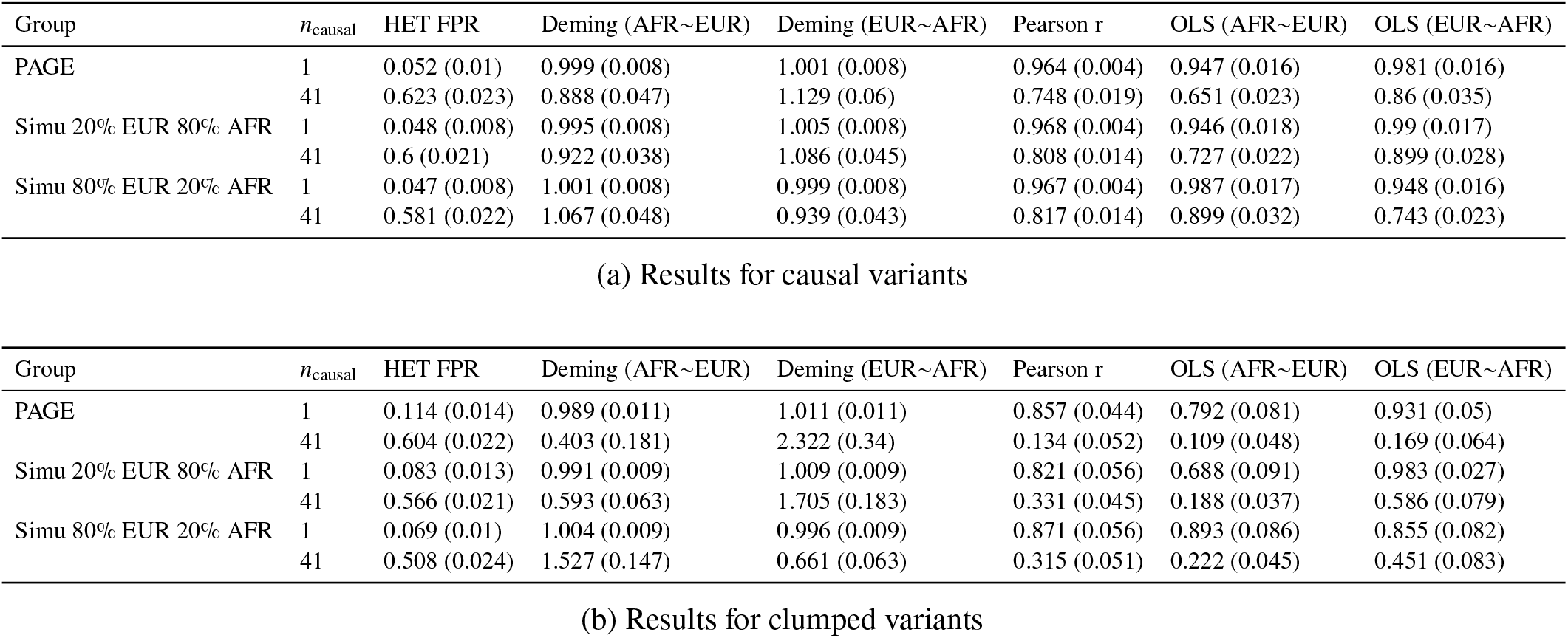
Simulation results in PAGE data set and simulated genotype data sets with varying ancestry proportion (Figure S8). We report numerical results for the 4 metrics in genotype data sets with varying ancestry proportion. For each data set, we varied *n*_causal_ = 1, 41 and fixed the heritability at 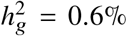. We report the mean and standard errors for each metric. Standard errors (displayed in parentheses) are based on 100 random sub-samplings with each sample consists of 500 SNPs. See details for simulation in Figure S8.

## References

[1] Genevieve L Wojcik, Mariaelisa Graff, Katherine K Nishimura, Ran Tao, Jeffrey Haessler, Christopher R Gignoux, Heather M Highland, Yesha M Patel, Elena P Sorokin, Christy L Avery, et al. Genetic analyses of diverse populations improves discovery for complex traits. Nature, 570(7762):514–518, 2019.

[2] Clare Bycroft, Colin Freeman, Desislava Petkova, Gavin Band, Lloyd T Elliott, Kevin Sharp, Allan Motyer, Damjan Vukcevic, Olivier Delaneau, Jared O’Connell, et al. The uk biobank resource with deep phenotyping and genomic data. Nature, 562(7726):203–209, 2018.

[3] Andrea H Ramirez, Lina Sulieman, David J Schlueter, Alese Halvorson, Jun Qian, Francis Ratsimbazafy, Roxana Loperena, Kelsey Mayo, Melissa Basford, Nicole Deflaux, et al. The all of us research program: data quality, utility, and diversity. medRxiv, 2020.

[4] Wei Zhou, Global Biobank Meta analysis Initiative, et al. Global biobank meta-analysis initiative: Powering genetic discovery across human diseases. medRxiv, 2021.

[5] Brielin C Brown, Chun Jimmie Ye, Alkes L Price, Noah Zaitlen, Asian Genetic Epidemiology Network Type 2 Diabetes Consortium, et al. Transethnic genetic-correlation estimates from summary statistics. The American Journal of Human Genetics, 99(1):76–88, 2016.

[6] Kevin J Galinsky, Yakir A Reshef, Hilary K Finucane, Po-Ru Loh, Noah Zaitlen, Nick J Patterson, Brielin C Brown, and Alkes L Price. Estimating cross-population genetic correlations of causal effect sizes. Genetic epidemiology, 43(2):180–188, 2019.

[7] Huwenbo Shi, Kathryn S Burch, Ruth Johnson, Malika K Freund, Gleb Kichaev, Nicholas Mancuso, Astrid M Manuel, Natalie Dong, and Bogdan Pasaniuc. Localizing components of shared transethnic genetic architecture of complex traits from gwas summary data. The American Journal of Human Genetics, 106(6):805–817, 2020.

[8] Huwenbo Shi, Steven Gazal, Masahiro Kanai, Evan M Koch, Armin P Schoech, Katherine M Siewert, Samuel S Kim, Yang Luo, Tiffany Amariuta, Hailiang Huang, et al. Population-specific causal disease effect sizes in functionally important regions impacted by selection. Nature communications, 12(1):1–15, 2021.

[9] Masahiro Kanai, Jacob C Ulirsch, Juha Karjalainen, Mitja Kurki, Konrad J Karczewski, Eric Fauman, Qingbo S Wang, Hannah Jacobs, François Aguet, Kristin G Ardlie, et al. Insights from complex trait fine-mapping across diverse populations. medRxiv, 2021.

[10] Deepti Gurdasani, Inês Barroso, Eleftheria Zeggini, and Manjinder S Sandhu. Genomics of disease risk in globally diverse populations. Nature Reviews Genetics, 20(9):520–535, 2019.

[11] Giorgio Sirugo, Scott M Williams, and Sarah A Tishkoff. The missing diversity in human genetic studies. Cell, 177(1):26–31, 2019.

[12] Alicia R Martin, Masahiro Kanai, Yoichiro Kamatani, Yukinori Okada, Benjamin M Neale, and Mark J Daly. Clinical use of current polygenic risk scores may exacerbate health disparities. Nature genetics, 51(4): 584–591, 2019.

[13] Ying Wang, Jing Guo, Guiyan Ni, Jian Yang, Peter M Visscher, and Loic Yengo. Theoretical and empirical quantification of the accuracy of polygenic scores in ancestry divergent populations. Nature communications, 11(1):1–9, 2020.

[14] Urko M Marigorta and Arcadi Navarro. High trans-ethnic replicability of gwas results implies common causal variants. PLoS genetics, 9(6):e1003566, 2013.

[15] Roshni A Patel, Shaila A Musharoff, Jeffrey P Spence, Harold Pimentel, Catherine Tcheandjieu, Hakhamanesh Mostafavi, Nasa Sinnott-Armstrong, Shoa L Clarke, Courtney J Smith, Peter P Durda, et al. Effect sizes of causal variants for gene expression and complex traits differ between populations. bioRxiv, 2021.

[16] Na Cai, Joana A Revez, Mark J Adams, Till FM Andlauer, Gerome Breen, Enda M Byrne, Toni-Kim Clarke, Andreas J Forstner, Hans J Grabe, Steven P Hamilton, et al. Minimal phenotyping yields genome-wide association signals of low specificity for major depression. Nature genetics, 52(4):437–447, 2020.

[17] Michael F Seldin, Bogdan Pasaniuc, and Alkes L Price. New approaches to disease mapping in admixed populations. Nature Reviews Genetics, 12(8):523–528, 2011.

[18] Melinda C Mills and Charles Rahal. The gwas diversity monitor tracks diversity by disease in real time. Nature genetics, 52(3):242–243, 2020.

[19] Elizabeth G Atkinson, Adam X Maihofer, Masahiro Kanai, Alicia R Martin, Konrad J Karczewski, Marcos L Santoro, Jacob C Ulirsch, Yoichiro Kamatani, Yukinori Okada, Hilary K Finucane, et al. Tractor uses local ancestry to enable the inclusion of admixed individuals in gwas and to boost power. Nature Genetics, 53(2): 195–204, 2021.

[20] Kangcheng Hou, Arjun Bhattacharya, Rachel Mester, Kathryn S Burch, and Bogdan Pasaniuc. On powerful gwas in admixed populations. Nature genetics, 53(12):1631–1633, 2021.

[21] Bárbara D Bitarello and Iain Mathieson. Polygenic scores for height in admixed populations. G3: Genes, Genomes, Genetics, 10(11):4027–4036, 2020.

[22] Davide Marnetto, Katri Pärna, Kristi Läll, Ludovica Molinaro, Francesco Montinaro, Toomas Haller, Mait Metspalu, Reedik Mägi, Krista Fischer, and Luca Pagani. Ancestry deconvolution and partial polygenic score can improve susceptibility predictions in recently admixed individuals. Nature communications, 11(1):1–9, 2020.

[23] Amy R Bentley, Guanjie Chen, Daniel Shriner, Ayo P Doumatey, Jie Zhou, Hanxia Huang, James C Mullikin, Robert W Blakesley, Nancy F Hansen, Gerard G Bouffard, et al. Gene-based sequencing identifies lipidinfluencing variants with ethnicity-specific effects in african americans. PLoS genetics, 10(3):e1004190, 2014.

[24] Farid Rajabli, Briseida E Feliciano, Katrina Celis, Kara L Hamilton-Nelson, Patrice L Whitehead, Larry D Adams, Parker L Bussies, Clara P Manrique, Alejandra Rodriguez, Vanessa Rodriguez, et al. Ancestral origin of apoe s4 alzheimer disease risk in puerto rican and african american populations. PLoS genetics, 14(12): e1007791, 2018.

[25] Elizabeth E Blue, Andréa RVR Horimoto, Shubhabrata Mukherjee, Ellen M Wijsman, and Timothy A Thornton. Local ancestry at apoe modifies alzheimer’s disease risk in caribbean hispanics. Alzheimer’s & Dementia, 15(12):1524–1532, 2019.

[26] Michel Satya Naslavsky, Claudia K Suemoto, Luciano Abreu Brito, Marilia Oliveira Scliar, Renata Eloah Ferretti-Rebustini, Roberta Diehl Rodriguez, Renata Paraizo Leite, Nathalia Matta Araujo, Victor Borda, Eduardo Tarazona-Santos, et al. Global and local ancestry modulate apoe association with alzheimer’s neuropathology and cognitive outcomes in an admixed sample. medRxiv, 2022.

[27] Jian Yang, S Hong Lee, Michael E Goddard, and Peter M Visscher. Gcta: a tool for genome-wide complex trait analysis. The American Journal of Human Genetics, 88(1):76–82, 2011.

[28] Saori Sakaue, Masahiro Kanai, Yosuke Tanigawa, Juha Karjalainen, Mitja Kurki, Seizo Koshiba, Akira Narita, Takahiro Konuma, Kenichi Yamamoto, Masato Akiyama, et al. A cross-population atlas of genetic associations for 220 human phenotypes. Nature genetics, 53(10):1415–1424, 2021.

[29] Jian Zeng, Ronald De Vlaming, Yang Wu, Matthew R Robinson, Luke R Lloyd-Jones, Loic Yengo, Chloe X Yap, Angli Xue, Julia Sidorenko, Allan F McRae, et al. Signatures of negative selection in the genetic architecture of human complex traits. Nature genetics, 50(5):746–753, 2018.

[30] Armin P Schoech, Daniel M Jordan, Po-Ru Loh, Steven Gazal, Luke J O’Connor, Daniel J Balick, Pier F Palamara, Hilary K Finucane, Shamil R Sunyaev, and Alkes L Price. Quantification of frequency-dependent genetic architectures in 25 uk biobank traits reveals action of negative selection. Nature communications, 10 (1):1–10, 2019.

[31] Yan Zhang, Guanghao Qi, Ju-Hyun Park, and Nilanjan Chatterjee. Estimation of complex effect-size distributions using summary-level statistics from genome-wide association studies across 32 complex traits. Nature genetics, 50(9):1318–1326, 2018.

[32] International HapMap 3 Consortium et al. Integrating common and rare genetic variation in diverse human populations. Nature, 467(7311):52, 2010.

[33] Doug Speed and David J Balding. Sumher better estimates the snp heritability of complex traits from summary statistics. Nature genetics, 51(2):277–284, 2019.

[34] William Edwards Deming. Statistical adjustment of data. 1943.

[35] Bogdan Pasaniuc, Noah Zaitlen, Guillaume Lettre, Gary K Chen, Arti Tandon, WH Linda Kao, Ingo Ruczinski, Myriam Fornage, David S Siscovick, Xiaofeng Zhu, et al. Enhanced statistical tests for gwas in admixed populations: assessment using african americans from care and a breast cancer consortium. PLoS genetics, 7 (4):e1001371, 2011.

[36] Chani J Hodonsky, Antoine R Baldassari, Stephanie A Bien, Laura M Raffield, Heather M Highland, Colleen M Sitlani, Genevieve L Wojcik, Ran Tao, Marielisa Graff, Weihong Tang, et al. Ancestry-specific associations identified in genome-wide combined-phenotype study of red blood cell traits emphasize benefits of diversity in genomics. BMC genomics, 21(1):1–14, 2020.

[37] Po-Ru Loh, Gaurav Bhatia, Alexander Gusev, Hilary K Finucane, Brendan K Bulik-Sullivan, Samuela J Pollack, Teresa R de Candia, Sang Hong Lee, Naomi R Wray, Kenneth S Kendler, et al. Contrasting genetic architectures of schizophrenia and other complex diseases using fast variance-components analysis. Nature genetics, 47(12):1385–1392, 2015.

[38] Ruth Johnson, Kathryn S Burch, Kangcheng Hou, Mario Paciuc, Bogdan Pasaniuc, and Sriram Sankararaman. Estimation of regional polygenicity from gwas provides insights into the genetic architecture of complex traits. PLoS computational biology, 17(10):e1009483, 2021.

[39] Bogdan Pasaniuc, Sriram Sankararaman, Dara G Torgerson, Christopher Gignoux, Noah Zaitlen, Celeste Eng, William Rodriguez-Cintron, Rocio Chapela, Jean G Ford, Pedro C Avila, et al. Analysis of latino populations from gala and mec studies reveals genomic loci with biased local ancestry estimation. Bioinformatics, 29(11): 1407–1415, 2013.

[40] Katarzyna Bryc, Eric Y Durand, J Michael Macpherson, David Reich, and Joanna L Mountain. The genetic ancestry of african americans, latinos, and european americans across the united states. The American Journal of Human Genetics, 96(1):37–53, 2015.

[41] Qiongshi Lu, Boyang Li, Derek Ou, Margret Erlendsdottir, Ryan L Powles, Tony Jiang, Yiming Hu, David Chang, Chentian Jin, Wei Dai, et al. A powerful approach to estimating annotation-stratified genetic covariance via gwas summary statistics. The American Journal of Human Genetics, 101(6):939–964, 2017.

[42] Jian Yang, Beben Benyamin, Brian P McEvoy, Scott Gordon, Anjali K Henders, Dale R Nyholt, Pamela A Madden, Andrew C Heath, Nicholas G Martin, Grant W Montgomery, et al. Common snps explain a large proportion of the heritability for human height. Nature genetics, 42(7):565–569, 2010.

[43] Steven Gazal, Hilary K Finucane, Nicholas A Furlotte, Po-Ru Loh, Pier Francesco Palamara, Xuanyao Liu, Armin Schoech, Brendan Bulik-Sullivan, Benjamin M Neale, Alexander Gusev, et al. Linkage disequilibrium– dependent architecture of human complex traits shows action of negative selection. Nature genetics, 49(10): 1421–1427, 2017.

[44] Steven Gazal, Carla Marquez-Luna, Hilary K Finucane, and Alkes L Price. Reconciling s-ldsc and ldak functional enrichment estimates. Nature genetics, 51(8):1202–1204, 2019.

[45] Kangcheng Hou, Kathryn S Burch, Arunabha Majumdar, Huwenbo Shi, Nicholas Mancuso, Yue Wu, Sriram Sankararaman, and Bogdan Pasaniuc. Accurate estimation of snp-heritability from biobank-scale data irrespective of genetic architecture. Nature genetics, 51(8):1244–1251, 2019.

[46] Kristian Linnet. Performance of deming regression analysis in case of misspecified analytical error ratio in method comparison studies. Clinical chemistry, 44(5):1024–1031, 1998.

[47] Alec M Chiu, Erin K Molloy, Zilong Tan, Ameet Talwalkar, and Sriram Sankararaman. Inferring population structure in biobank-scale genomic data. bioRxiv, 2021.

[48] Po-Ru Loh, Petr Danecek, Pier Francesco Palamara, Christian Fuchsberger, Yakir A Reshef, Hilary K Finucane, Sebastian Schoenherr, Lukas Forer, Shane McCarthy, Goncalo R Abecasis, et al. Reference-based phasing using the haplotype reference consortium panel. Nature genetics, 48(11):1443–1448, 2016.

[49] Brian K Maples, Simon Gravel, Eimear E Kenny, and Carlos D Bustamante. Rfmix: a discriminative modeling approach for rapid and robust local-ancestry inference. The American Journal of Human Genetics, 93(2): 278–288, 2013.

[50] 1000 Genomes Project Consortium et al. A global reference for human genetic variation. Nature, 526(7571): 68, 2015.

[51] Jianqi Zhang and Daniel O Stram. The role of local ancestry adjustment in association studies using admixed populations. Genetic epidemiology, 38(6):502–515, 2014.

[52] Nicole R Gay, Michael Gloudemans, Margaret L Antonio, Nathan S Abell, Brunilda Balliu, YoSon Park, Alicia R Martin, Shaila Musharoff, Abhiram S Rao, François Aguet, et al. Impact of admixture and ancestry on eqtl analysis and gwas colocalization in gtex. Genome biology, 21(1):1–20, 2020.

[53] Antonio F Pardiñas, Peter Holmans, Andrew J Pocklington, Valentina Escott-Price, Stephan Ripke, Noa Carrera, Sophie E Legge, Sophie Bishop, Darren Cameron, Marian L Hamshere, et al. Common schizophrenia alleles are enriched in mutation-intolerant genes and in regions under strong background selection. Nature genetics, 50(3):381–389, 2018.

[54] Armin P Schoech, Omer Weissbrod, Luke J O’Connor, Nick Patterson, Huwenbo Shi, Yakir Reshef, and Alkes L Price. Negative short-range genomic autocorrelation of causal effects on human complex traits. bioRxiv, 2020.

[55] David Reich, Michael A Nalls, WH Linda Kao, Ermeg L Akylbekova, Arti Tandon, Nick Patterson, James Mullikin, Wen-Chi Hsueh, Ching-Yu Cheng, Josef Coresh, et al. Reduced neutrophil count in people of african descent is due to a regulatory variant in the duffy antigen receptor for chemokines gene. PLoS genetics, 5(1): e1000360, 2009.

[56] Alex P Reiner, Sandra Beleza, Nora Franceschini, Paul L Auer, Jennifer G Robinson, Charles Kooperberg, Ulrike Peters, and Hua Tang. Genome-wide association and population genetic analysis of c-reactive protein in african american and hispanic american women. The American Journal of Human Genetics, 91(3):502–512, 2012.

[57] Ani Manichaikul, Josyf C Mychaleckyj, Stephen S Rich, Kathy Daly, Michèle Sale, and Wei-Min Chen. Robust relationship inference in genome-wide association studies. Bioinformatics, 26(22):2867–2873, 2010.

[58] Christopher C Chang, Carson C Chow, Laurent CAM Tellier, Shashaank Vattikuti, Shaun M Purcell, and James J Lee. Second-generation plink: rising to the challenge of larger and richer datasets. Gigascience, 4(1): s13742–015, 2015.

[59] Gao Wang, Abhishek Sarkar, Peter Carbonetto, and Matthew Stephens. A simple new approach to variable selection in regression, with application to genetic fine mapping. Journal of the Royal Statistical Society: Series B (Statistical Methodology), 82(5):1273–1300, 2020.

